# A limbic-predominant amnestic neurodegenerative syndrome associated with TDP-43 pathology

**DOI:** 10.1101/2023.11.19.23298314

**Authors:** Nick Corriveau-Lecavalier, Hugo Botha, Jonathan Graff-Radford, Aaron R. Switzer, Scott A. Przybelski, Heather J. Wiste, Melissa E. Murray, R. Ross Reichard, Dennis W. Dickson, Aivi T. Nguyen, Vijay K. Ramanan, Stuart J. McCarter, Bradley F. Boeve, Mary M. Machulda, Julie A. Fields, Nikki H. Stricker, Peter T. Nelson, Michel J. Grothe, David S. Knopman, Val J. Lowe, Ronald C. Petersen, Clifford R. Jack, David T. Jones

## Abstract

Limbic-predominant age-related TDP-43 encephalopathy (LATE) is a neuropathologically-defined disease that affects 40% of persons in advanced age, but its associated neurological syndrome is not defined. LATE neuropathological changes (LATE-NC) are frequently comorbid with Alzheimer’s disease neuropathologic changes (ADNC). When seen in isolation, LATE-NC have been associated with a predominantly amnestic profile and slow clinical progression. We propose a set of clinical criteria for a limbic-predominant amnestic neurodegenerative syndrome (LANS) that is highly associated with LATE-NC but also other pathologic entities. The LANS criteria incorporate core, standard and advanced features that are measurable *in vivo*, including older age at evaluation, mild clinical syndrome, disproportionate hippocampal atrophy, impaired semantic memory, limbic hypometabolism, absence of neocortical degenerative patterns and low likelihood of neocortical tau, with degrees of certainty (highest, high, moderate, low). We operationalized this set of criteria using clinical, imaging and biomarker data to validate its associations with clinical and pathologic outcomes. We screened autopsied patients from Mayo Clinic (n = 922) and ADNI (n = 93) cohorts and applied the LANS criteria to those with an antemortem predominant amnestic syndrome (Mayo, *n* = 165; ADNI, *n* = 53). ADNC, ADNC/LATE-NC and LATE-NC accounted for 35%, 37% and 4% of cases in the Mayo cohort, respectively, and 30%, 22%, and 9% of cases in the ADNI cohort, respectively. The LANS criteria effectively categorized these cases, with ADNC having the lowest LANS likelihoods, LATE-NC patients having the highest likelihoods, and ADNC/LATE-NC patients having intermediate likelihoods. A logistic regression model using the LANS features as predictors of LATE-NC achieved a balanced accuracy of 74.6% in the Mayo cohort, and out-of-sample predictions in the ADNI cohort achieved a balanced accuracy of 73.3%. Patients with high LANS likelihoods had a milder and slower clinical course and more severe temporo-limbic degeneration compared to those with low likelihoods. Stratifying ADNC/LATE-NC patients from the Mayo cohort according to their LANS likelihood revealed that those with higher likelihoods had more temporo-limbic degeneration and a slower rate of cognitive decline, and those with lower likelihoods had more lateral temporo-parietal degeneration and a faster rate of cognitive decline. The implementation of LANS criteria has implications to disambiguate the different driving etiologies of progressive amnestic presentations in older age and guide prognosis, treatment, and clinical trials. The development of *in vivo* biomarkers specific to TDP-43 pathology are needed to refine molecular associations between LANS and LATE-NC and precise antemortem diagnoses of LATE.

## Introduction

Limbic-predominant age-related TDP-43 encephalopathy (LATE) is defined by neuropathological changes (LATE-NC)(1) that are distinct from fronto-temporal lobar degeneration (FTLD) neuropathology, or other disorders with TDP-43(1–4). The stereotypical pattern of LATE-NC development in the brain first involves the amygdala, followed by the hippocampus, and then the middle frontal gyrus(5–9). Prevalence increases with older age, and it is estimated that ∼40% of autopsied brains beyond age 85 years have LATE-NC(10,11). LATE-NC is frequently co-morbid with other neuropathologies including amyloid plaques and tau tangles, i.e. Alzheimer’s disease neuropathological changes (ADNC), with which LATE-NC partially share pathogenic mechanisms(9,12–15).

Several efforts based on retrospective clinicopathological studies have aimed to characterize the clinical profile of individuals who had LATE-NC at autopsy, and to disambiguate this profile from those who had ADNC without LATE-NC. The most common finding across studies is that memory impairment dominates the clinical profile of individuals with LATE-NC while cognitive functions functionally associated with neocortical areas (e.g., visuospatial processing) are relatively preserved(6,16–18). Studies that evaluated the longitudinal trajectory of cognitive impairment in LATE-NC consistently found a milder and slower decline of memory and global function relative to ADNC, and a steeper rate of decline in patients with evidence for both diseases, i.e., ADNC/LATE-NC(16,19–21). Evidence of non-memory impairment has also been documented in LATE-NC and mostly pertains to semantic knowledge as measured through confrontation naming, verbal fluency, and knowledge of famous faces and events(16,22,23). However, these studies have not directly compared patients with LATE-NC versus ADNC and/or did not always account for global severity of impairment. It is thus unclear whether lower scores on tasks of semantic knowledge are explained by global severity as opposed to a genuine specificity to LATE-NC. Overall, evidence supports a clinical profile characterized by a relatively isolated amnestic syndrome with an indolent progression in patients with LATE-NC relative to ADNC, in addition to semantic knowledge impairment, although less documented.

*In vivo* neuroimaging markers of autopsy-confirmed LATE-NC have been described. The hippocampus is a known locus of LATE-NC pathology. For example, MRI studies have repeatedly found smaller hippocampal volume and faster rates of hippocampal atrophy associated with LATE-NC when accounting for the extent of ADNC(14,24–26). Hippocampal sclerosis (severe cell loss and gliosis in the hippocampal formation, often with accentuated atrophy) is a frequent finding at autopsy associated with LATE-NC(1,14,27). In fact, studies have described patients with a dense amnestic syndrome who were either tau-negative on PET imaging and/or had TDP-43 related hippocampal sclerosis with minimal ADNC at autopsy(28,29). This strongly supports the hypothesis that LATE-NC is an independent driver of hippocampal atrophy that is sufficient to cause a progressive amnestic syndrome.

FDG-PET has also been proven useful for delineating patterns of involvement associated with LATE-NC and has the potential to objectively index impaired limbic function in the setting of preserved neocortical function. Botha et al.(28) first developed the inferior-to-medial temporal (IMT) ratio as a marker of TDP-43 related hippocampal sclerosis in tau-negative amnestic dementia, where such patients exhibit prominent medial temporal lobe hypometabolism relative to the lateral temporal lobe, while the opposite pattern is typically associated with ADNC. Buciuc et al. subsequently showed that this marker is specific and sensitive to advanced LATE-NC regardless of hippocampal sclerosis status(30). A recent study by Grothe et al.(31) showed that autopsy-confirmed cases of LATE-NC exhibit prominent temporo-limbic hypometabolism compared to those with ADNC, and that independent patients with a clinical diagnosis of Alzheimer’s disease dementia showing this “LATE-NC-like” pattern had an older age at evaluation, a memory-dominant cognitive impairment profile, a slower clinical course and lower Alzheimer’s disease biomarkers levels. These findings position temporo-limbic-predominant hypometabolism, in addition to disproportionate hippocampal atrophy on MRI, as a promising candidate for the *in vivo* identification of individuals with predominant limbic degeneration highly associated with underlying LATE-NC and for distinguishing such patients from those with neocortical degeneration and ADNC as the primary driver of their symptoms.

Despite the significant advances in identifying *in vivo* features indicative of LATE-NC, clinically applicable criteria of a predominant amnestic syndrome driven by a pattern of limbic-predominant degeneration that is highly associated with TDP-43 pathology do not currently exist. The implementation of such criteria is critical because an aspirational goal of clinical and research endeavors on LATE-NC is to define a constellation of clinical symptoms that is driven by LATE-NC, i.e., LATE. This requires both a set of clinical criteria associated with limbic degeneration and *in vivo* diagnostic biomarkers of TDP-43. The current investigation has the former as its primary objective. The *in vivo* identification of patients with a high likelihood of limbic degeneration due to underlying LATE-NC as the primary driver of their symptoms has relevance for symptom management and prognosis, which are thought to differ from other amnestic syndromes associated with other common primary neuropathologies such as ADNC. The detection of non-AD causes for predominant amnestic symptoms is also highly relevant in this era of emerging disease-modifying therapies to prevent patients from being inadvertently treated with inappropriate therapies(32). The identification of patients with a high likelihood of LATE-NC is also important for refining the clinical phenotype associated with predominant limbic degeneration and which will be critical for developing clinical trials aimed at TDP-43 pathology.

We propose a set of clinical criteria for a limbic-predominant amnestic neurodegenerative syndrome (LANS). LANS is defined as a degenerative neurologic syndrome in that it refers to a set of clinical signs and symptoms associated with a requisite functional neuroanatomic localization, i.e., the limbic system. All available relevant information (e.g., history, physical, neuropsychologic evaluation, imaging, and fluids) should be used to inform a standard neurologic evaluation of this and related degenerative neurologic syndromes across a spectrum of functional neuroanatomic localizations(33). While the definition of LANS is agnostic to molecular pathology, this syndrome is highly associated with LATE-NC but also other less common neuropathologic entities selectively targeting the limbic system, for instance limbic-predominant forms of Alzheimer’s disease or argyrophilic grain disease (AGD). Given that LANS does not perfectly map onto a single underlying pathology and that there is currently no clinically applicable *in vivo* biomarker of TDP-43, it does not meet the definition of a clinicopathologic entity(34). If an accepted TDP-43 biomarker is developed it could be combined with LANS to define a clinicopathologic entity like LATE. The current investigation focuses on LANS associations with LATE-NC, but qualitative data on other possible causes of LANS are provided in Supplemental Materials and are referenced in-text where appropriate. The LANS criteria includes core, standard, and advanced criteria that can be measured *in vivo* along with levels of certainty (described in Box 1). We operationalized this set of criteria based on clinical, imaging, and biomarker features. It is important to note that the LANS clinical criteria and associated likelihoods are meant to guide clinical decision making in practice (see Box 2 for an example of the clinical application of the LANS criteria). Our operationalization of the proposed LANS criteria only applies to the current study; we use this operationalization to support the construct validity of the proposed LANS criteria by retrospectively applying it to a cohort of autopsied patients from the Mayo Clinic and ADNI cohorts with an antemortem history of a predominant and progressive amnestic syndrome. It thus serves a validation purpose in the context of the current and future studies and is not meant to be concretely applied in clinical practice.

## Methods

### Participants

Two clinicopathological cohorts were used for this study. The primary cohort came from the Mayo Clinic Study of Aging (MCSA) and Alzheimer’s Disease Research Center (ADRC) research programs from Mayo Clinic Rochester (now referred to as “Mayo cohort”). The second cohort came from the Alzheimer’s Disease Neuroimaging Initiative (ADNI cohort). We screened all autopsied patients from these two cohorts (Mayo cohort, *n* = 922, ADNI cohort, *n* = 93) and included those with an antemortem history of a predominant and progressive amnestic syndrome. This was defined by a clinical diagnosis of AD dementia or single- or multi-domain amnestic mild cognitive impairment (aMCI) at baseline according to widely accepted criteria(35–37). We additionally excluded patients with insufficient pathology data, i.e., missing information about amyloid plaques, tau and Lewy Bodies, and LATE-NC. The primary analyses only considered patients with a pathological diagnosis of ADNC, LATE-NC, or comorbid ADNC/LATE-NC. This resulted in a final sample of 165 patients from the Mayo cohort and 53 patients from the ADNI cohort. The workflow of patient inclusion is found in Supplementary Figure 1. Analyses considering all primary neuropathological diagnoses associated with a progressive and predominant amnestic syndrome were conducted separately and are in Supplementary Materials (referenced in-text where appropriate). Data from 112 age- and sex-matched cognitively unimpaired (CU) controls from the MCSA were collected for imaging comparisons purposes (MRI, FDG-PET) in the Mayo cohort. CUs had to be amyloid- and tau-negative based on PET imaging and have MRI and FDG-PET imaging available for inclusion in the study.

Patients and/or their legal representative provided written consent for their data to be used for research purposes. This study met HIPAA guidelines and was approved by the Mayo Clinic Institutional Review Board.

### Neuropathological assessment

Assessments for the Mayo and ADNI cohorts were performed by experienced neuropathologists in accordance with current diagnostic protocols(38) and are described in Supplementary Materials. ADNC was diagnosed according to the ABC ranking score(39) which includes the Thal staging of amyloid plaques, Braak staging of neurofibrillary tangles(40) and the density measurement of neuritic plaques(38). A stage of Braak IV, V or VI in the absence of significant amyloidosis (<<u>2</u> Thal stage) was classified as primary age-related tauopathy (PART)(41). In the Mayo cohort, TDP-43 type-A was defined as TDP-43 immunoreactive neuronal cytoplasmic inclusions, dystrophic neurites and neuronal intranuclear inclusions in vulnerable cortical and subcortical areas. TDP-43 type B had predominantly neuronal cytoplasmic inclusions(42). Similar procedures were applied in ADNI (see https://adni.loni.usc.edu/methods/neuropath-methods/). TDP-43 staging was classified as FTLD-TDP-43 related or not (i.e., LATE-NC) based on its spatial distribution. LATE-NC staging was done in 40/90 of patients with confirmed TDP-43 from the Mayo cohort and all patients from the ADNI cohort. Lewy Body Disease (LBD) was staged according to published criteria(43) and significant burden was considered when pathology was documented in limbic and/or neocortical areas. Corticobasal degeneration (CBD) was diagnosed by the presence of cortical and subcortical neuronal and glial lesions (i.e., astrocytic plaques) and thread-like processes in gray and white matter(44). Progressive supranuclear palsy (PSP) staging was performed according to published criteria(45). CBD and PSP were categorized as “FTLD-tau”. A diagnosis of AGD was made if there were silver and tau-positive spindle-shaped lesions, coiled bodies and balloon neurons in trans-entorhinal and entorhinal cortex, amygdala or cingulate gyrus(46). The presence of other pathologies including hippocampal sclerosis and vascular disease (cerebral amyloid angiopathy, infarcts and lacunes, microbleeds, hemorrhages, arteriolosclerosis) was also assessed.

### Imaging acquisition and processing

Acquisition protocols for MRI and PET images are in Supplementary Materials. Regional MRI and FDG-PET data were generated for both the Mayo and ADNI cohorts using in-house processing pipelines from Mayo Clinic using Statistical Parameter Mapping 12 (SPM12). PET images were co-registered to their corresponding MRI image. All images were normalized into the Mayo Clinic Adult Lifespan Template (MCALT) and smoothed with a 6-millimeter full width at half-maximum smoothing kernel. FDG-PET images were normalized to the pons to yield regional standard uptake value ratios (SUVR). The FDG-PET IMT ratio was derived by dividing SUVR values from the inferior temporal lobe by the size-weighted sum of SUVR values from the amygdala and hippocampus(28,30), where higher values are indicative of LATE (thresholding procedures are described in the Statistical analyses section).

Hippocampal volume calculation is described in separate publications(47,48). Briefly, it was corrected for intracranial volume by calculating the residuals from a linear regression based on a sex-specific formula. This is similar to the approach from Jack et al.(48), except for the use of SPM12 instead of FreeSurfer and a different CU sample as described in Stricker et al.(47). This measure was combined across hemispheres.

Different methods were used to determine abnormality thresholds for global FDG-PET, amyloid-PET and tau-PET SUVR meta-regions of interest (meta-ROIs) across the Mayo and ADNI cohorts due to differences in processing pipelines, radiotracers for amyloid-PET and cohorts studied. This was done only for global measures, whereas regional data were generated using the same pipeline as described above. Briefly, global FDG-PET (normalized to the pons), amyloid-PET and tau-PET (normalized to the cerebellar crus) were derived from established meta-ROIs(49–51). Images from the Mayo cohort processed with the pipeline described above were used, and abnormality thresholds set at ≤1.47 (FDG-PET), ≥1.48 (amyloid-PET; PiB) and ≥1.29 (tau-PET; Flortaucipir) according to published methods(49,52). Images from the ADNI cohort processed in Berkeley (CA, USA) using published methods(53) were used. Abnormality thresholds were set at ≤1.21 (FDG-PET), ≥1.11 (amyloid-PET; Florbetapir), ≥1.0818 (amyloid-PET; Florbetaben), and ≥1.29 (tau-PET; Flortaucipir) according to published methods(50,53–55). Amyloid-PET values from both cohorts were transformed into centiloids for descriptive purposes.

### Fluid biomarkers

CSF samples from both the Mayo and ADNI cohorts were analyzed using published protocols(56,57). Briefly, samples were analyzed using Elecsys β-Amyloid (1–42) CSF, Total-Tau CSF, and Phospho-Tau (181P) CSF electrochemiluminescence immunoassays (Roche Diagnostics). Quality control procedures and technical limits were handled as previously described(56). Thresholds were set at ≤1026 pg/ml for Aβ42, ≥22 pg/ml for P-tau, and ≥0.023 for the Aβ42/P-tau ratio(56,58,59).

Procedures for the analysis and quality control of plasma assays for the Mayo cohort are described in a separate publication(60). Briefly, plasma phosphorylated-Tau 181 (pTau181) was measured with the Simoa^®^ pTau-181 Advantage V2 kit following instructions from the manufacturer and ran on a Quanterix HD-X analyzer (Quanterix, Lexington, MA, USA). The pTau181 threshold was set at <2.56 pg/ml according to previous research assessing the relationship between pTau181 and ADNC(60). Plasma assays in ADNI were collected and processed according to published methods(61,62) and were measured using an in-house assay as previously described(63). Plasma pTau181 was measured with Simoa HD-X instruments (Quanterix, Billerica, MA, USA) as described elsewhere(64). The pTau181 threshold was set at <17.7 pg/ml according to previous research assessing the relationship between pTau181 and Alzheimer’s disease biomarkers(64).

### Operationalization of the LANS criteria

The operationalization of the LANS criteria involves *in vivo* clinical, imaging and biomarker data. These operationalized criteria are not meant to be concretely applied to clinical practice: rather, their purpose is to help objectively validate our proposed LANS criteria. These operationalized criteria are as follows:

1. Age at evaluation (standard feature): age of ≥75 and older. Age at first visit was used.
2. Mild clinical syndrome (standard feature): diagnosis of mild cognitive impairment or mild dementia. In the context of this study, mild dementia was defined as a clinical diagnosis of Alzheimer’s-type dementia with a score ≤4 on the Clinical Dementia Rating Sum-of-Boxes (CDR-SB)(65). The score at the first visit was used.
3. Hippocampal atrophy out of proportion to syndrome severity (standard feature): hippocampal volume smaller than expected according to the CDR-SB score (see procedure in Statistical analyses). The hippocampal volume at the last MRI was used.
4. Mildly impaired semantic memory (standard feature): given the less well-established literature and the lack of gold standard measure to best assess this impairment in the context of LATE-NC, we decided to forgo the operationalization of this feature in this iteration of the LANS criteria. This feature is thus not included in the calculation of likelihoods in the context of the current study, but its use is encouraged in clinical practice using expert clinical judgement.
5. Limbic hypometabolism (advanced feature): above-threshold value on the FDG-PET IMT ratio, which is indicative of LATE-NC (see procedure in Statistical analyses). IMT ratio at the last FDG-PET was used.
6. Absence of neocortical degenerative disease pattern (advanced feature): above-threshold value on an established FDG-PET Alzheimer’s disease meta-ROI, which is indicative of an absence of the ADNC pattern. The score at the last FDG-PET was used. We used a meta-ROI indicative of underlying Alzheimer’s disease pathology in the context of this study, but this applies to any imaging biomarker indicative of neocortical degeneration.
7. Low likelihood of significant neocortical tau pathology (advanced feature): The rationale of this criterion is to reduce the likelihood that the clinical syndrome is primarily driven by neocortical pathology as seen in the multi-domain forms of Alzheimer’s disease. When multiple biomarkers were available, PET imaging was prioritized over fluid biomarkers. A negative amyloid-PET strongly reduces the likelihood of significant neocortical tau pathology and means that this criterion is met(66). A positive amyloid-PET scan should prompt further workup to assess the presence of neocortical tau pathology. The specification of “neocortical” tau as opposed to tau in general is to account for the limbic variant of Alzheimer’s disease wherein associated clinical and pathologic findings localize to the limbic system(67,68). Therefore, limbic Alzheimer’s disease, although less common than LATE-NC, qualifies for LANS. The visual assessment of neocortical tau can be guided by the approved FDA recommendations which includes the posterior lateral temporal lobe as the only temporal region to count towards neocortical tau positivity (see https://www.accessdata.fda.gov/drugsatfda_docs/label/2020/212123s000lbl.pdf). In regard to plasma biomarkers, current assays are mostly sensitive to amyloid pathology and current thresholds are not useful in determining limbic versus neocortical tau(69). The same applies to the CSF P-tau/Aβ ratio. Fluid biomarkers, in their current state of development, should only be used to rule out amyloidosis. If positive, further workup is recommended to investigate the presence of neocortical versus limbic distribution of tau, the key point being that a biomarker that is associated with an amyloid negative state is also considered a surrogate marker of an absence of neocortical tau(66). Conversely a biomarker that is associated with an amyloid positive state is not informative about neocortical tau for the purposes of the LANS criteria. This interpretation will apply even as novel plasma assays and associated methodologies more sensitive to tau staging are being developed and used in a clinical setting. For example, if a future blood-based measure is shown to be sensitive and specific for neocortical tau, then it could be used to meet this LANS criteria.

### Statistical analyses

Statistical analyses were performed using *R* version 4.2.3. One-way ANOVAs and chi-squared tests were used to assess between-group differences on demographic, clinical, and biomarker data, and post hoc analyses were performed when the omnibus test was significant.

We compared FDG-PET and MRI findings between ADNC, ADNC/LATE-NC, LATE-NC patients and CUs from the Mayo cohort in a pair-wise fashion using SPM12, resulting in *t* maps. We applied a false discovery rate correction to control for multiple comparisons at the voxel-level.

The procedure to assess disproportionate hippocampal atrophy according to clinical severity consisted of fitting a mixed linear model with CDR-SB as a predictor of hippocampal volume accounting for intra-individual change in the ADNI cohort. This was done including LATE-NC, ADNC and ADNC/LATE-NC patients only. We then calculated the scaled residuals between predicted and true values. We used a receiver operating characteristic (ROC) curve analysis on *Z* scored residuals to determine the optimal threshold for discriminating patients with LATE-NC (LATE and ADNC/LATE-NC) from those with ADNC based on the last MRI scan obtained. We applied this threshold in the ADNI cohort, and in the Mayo cohort after predicting the residuals using the model defined in ADNI. We also used a ROC curve analysis in the ADNI cohort to determine the FDG-PET IMT threshold that best discriminates individuals with LATE-NC (LATE and ADNC/LATE-NC) from those with ADNC based on the last FDG-PET scan obtained. We then applied this threshold in the ADNI and Mayo cohorts.

Counts for standard and advanced LANS features and likelihoods derived from the operationalized criteria were measured for each patient for each pathological diagnosis. Chi-squared analyses followed by post hoc tests were conducted to assess the distribution of pathological diagnoses within likelihoods and the distribution of likelihoods within pathological diagnoses. This was done while combining highest and high likelihoods within a single category. We then fit a mixed linear model with an interaction between time from baseline (in years) and likelihood group (highest, high, moderate, low) as predictor and CDR-SB as outcome measure accounting for intra-individual change to assess differences in clinical progression across likelihoods. We then assessed FDG-PET differences between likelihoods categories (highest/high, moderate, low) and CUs in pair-wise fashion as described above.

We fit a logistic regression model in the Mayo cohort to perform a binary classification of patients with LATE-NC (i.e., LATE-NC and ADNC/LATE-NC) versus ADNC using LANS features as input. Only the tau score was considered as a categorical variable (positive, negative) whereas other variables (age at examination, CDR-SB score, hippocampal volume, IMT ratio, FDG-PET meta-ROI) were considered as continuous. We then performed out-of-sample predictions in the ADNI cohort using the model fitted in the Mayo cohort.

We performed an exploratory analysis specifically in patients with ADNC/LATE-NC from Mayo Clinic on the premise that some of these patients may have LATE-NC as a primary pathology and ADNC as a secondary pathology, and vice-versa. We divided ADNC/LATE-NC patients into highest/high, moderate and low LANS likelihoods. We fit a mixed linear model with an interaction between time from baseline (in years) and group (ADNC, ADNC/LATE-NC highest/high, ADNC/LATE-NC moderate, ADNC/LATE-NC low, LATE-NC) as predictor and CDR-SB as outcome measure accounting for intra-individual change to assess differences in clinical progression across groups. We then assessed FDG-PET differences between ADNC/LATE-NC likelihood groups and CUs in a pair-wise fashion as described above.

## Data availability

Data from the Mayo Clinic Study of Aging and the Mayo Clinic Alzheimer’s Disease Research Center are available upon request (https://www.mayo.edu/research/centers-programs/alzheimers-disease-research-center/data-requests). ADNI data can be downloaded at https://adni.loni.usc.edu/. The MCALT can be found at: https://www.nitrc.org/projects/mcalt/.

## Results

### Sample characteristics

Demographic, clinical, and biomarker data at baseline for patients with ADNC, LATE-NC and ADNC/LATE-NC from the Mayo and ADNI cohorts are displayed in Table 1. The data for all pathological diagnoses are displayed in Supplementary Table 1. LATE-NC and ADNC/LATE-NC patients were older than ADNC patients at presentation and death in the Mayo cohort only. In both cohorts, there were more *APOE4* carriers in ADNC and ADNC/LATE-NC groups compared to LATE-NC, and ADNC and ADNC/LATE-NC groups had higher amyloid-PET centiloid values compared to LATE-NC. ADNC/LATE-NC patients had, on average, higher baseline CDR-SB scores than LATE-NC patients in the ADNI cohort only. There were no other differences.

**Table 1.** Demographic, clinical, and biomarker data of the Mayo and ADNI cohorts.

| Mayo cohort |  |  |  |  |
| --- | --- | --- | --- | --- |
|  | ADNC | ADNC/LATE-NC | LATE-NC | *P* |
| n | 75 | 81 | 9 | - |
| Age at first visit | 74.2 [63.7, 80.5] | 79 [72.2, 83.9] | 86.2 [83.6, 93.3] | < 0.001 |
| Age at death | 82.4 [71.1, 88.3] | 89.1 [83.9, 92.4] | 91.6 [88.7, 95.5] | < 0.001 |
| Sex (F, M) | 33, 42 | 36, 45 | 4, 5 | 0.8 |
| Education | 16 [12, 18] | 16 [12, 17] | 14 [13, 16] | 0.38 |
| CDR-SB | 1.5 [0.5, 3.5] | 1.5 [0.5, 3] | 0.5 [0.5, 1.25] | 0.39 |
| AD dementia | 27 | 30 | 1 | - |
| Amnestic MCI | 48 | 51 | 8 | - |
| APOE4 (+, -) | 47, 27 | 55, 24 | 1, 7 | 0.006 |
| Amyloid-PET centiloid | 110 [87.3, 129] | 110 [76.7, 122] | 9.75 [7.60, 13.4] | < 0.001 |
| AV1451 SUVR | 1.85 [1.72, 2.28] | 2.14 [1.40, 2.36] | 1.26* | - |
| ADNI cohort |  |  |  |  |
|  | ADNC | ADNC/LATE-NC | LATE-NC | *P* |
| n | 26 | 19 | 8 | - |
| Age at first visit | 75.8 [70.3, 81.1] | 77.7 [72.2, 85.5] | 74.9 [72.6, 81.7] | 0.4 |
| Age at death | 79 [76.2, 85.8] | 84 [79, 89] | 80.5 [79.8, 87] | 0.17 |
| Sex (F, M) | 9, 17 | 5, 14 | 1, 7 | 0.5 |
| Education | 16 [16, 18] | 16 [13.5, 17] | 16 [14.5, 18] | 0.28 |
| CDR-SB | 2 [1, 3.38] | 3.5 [2, 4.5] | 1 [0.875, 1.12] | 0.019 |
| AD dementia | 9 | 11 | 0 | - |
| Amnestic MCI | 17 | 8 | 8 | - |
| APOE4 (+, -) | 20, 6 | 13, 6 | 1, 7 | 0.004 |
| Amyloid-PET centiloid | 69.5 [50.2, 110] | 91 [81, 102] | -5 [-11.8, 0] | 0.0015 |
| AV1451 SUVR | NA | 1.52 [1.49, 1.55] | 1.08* | - |

\*One observation only. Values are expressed as counts or median and interquartile ranges. CDR-SB = Clinical Dementia Rating scale – Sum of Boxes; ADNC = Alzheimer’s disease neuropathological changes; LATE-NC = Limbic-predominant age-related TDP-43 encephalopathy neuropathological changes; MCI = Mild cognitive impairment; PET = Positron emission tomography; SUVR = Standard uptake value ratio.

The distribution of pathological diagnoses underlying a progressive and predominant neurodegenerative amnestic syndrome in the Mayo and ADNI cohorts is displayed in Figure 1. ADNC, ADNC/LATE-NC and LATE-NC accounted for 35%, 37% and 4% of cases in the Mayo cohort, respectively, and 30%, 22%, and 9% of cases in the ADNI cohort, respectively. Vascular pathologies (cerebral amyloid angiopathy, infarcts/lacunes, microbleeds, hemorrhages, arteriolosclerosis) were not considered in primary pathological diagnoses unless they were the only significant pathological feature (i.e., vascular dementia). The presence and severity of vascular pathologies across pathological diagnoses are listed in Supplementary Table 2.

**Figure 1.**
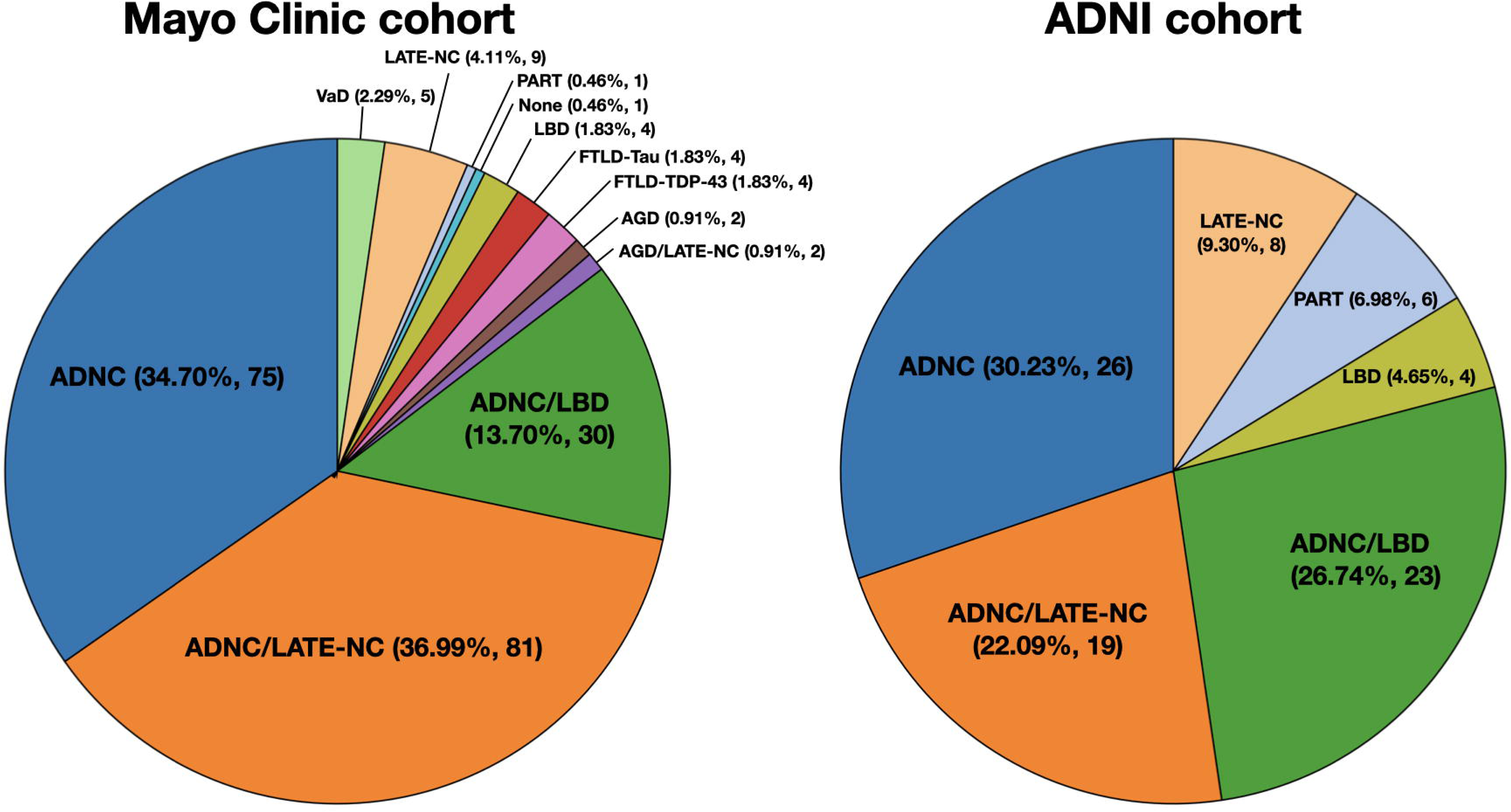
Primary neuropathological diagnoses associated with a predominant and progressive neurodegenerative amnestic syndrome in the Mayo and ADNI cohorts. ADNC = Alzheimer’s disease neuropathological changes; LATE-NC = Limbic-predominant age-associated TDP-43 encephalopathy neuropathological changes; LBD = Lewy body disease; AGD = Argyrophilic grain disease; FTLD = Fronto-temporal lobar degeneration; PART = Primary age-related tauopathy; VaD = Vascular disease.

### FDG-PET and MRI findings

FDG-PET and MRI contrasts comparing patients groups to CUs showed patterns of degeneration in lateral temporal and hippocampal areas in ADNC/LATE-NC patients and in lateral temporo-parietal and precuneus areas in ADNC. FDG-PET findings are displayed in Figure 2, and MRI findings are displayed in Supplementary Figure 2. Only one contrast between patient groups survived correction for multiple comparisons for both MRI and FDG-PET modalities. This contrast revealed that ADNC/LATE-NC had significantly more temporo-limbic degeneration involving the hippocampal, insular, temporopolar, middle frontal and orbitofrontal areas compared to ADNC patients. Comparisons involving LATE-NC patients failed to reveal significant differences at the group-level, likely owing to the small sample size.

**Figure 2.**
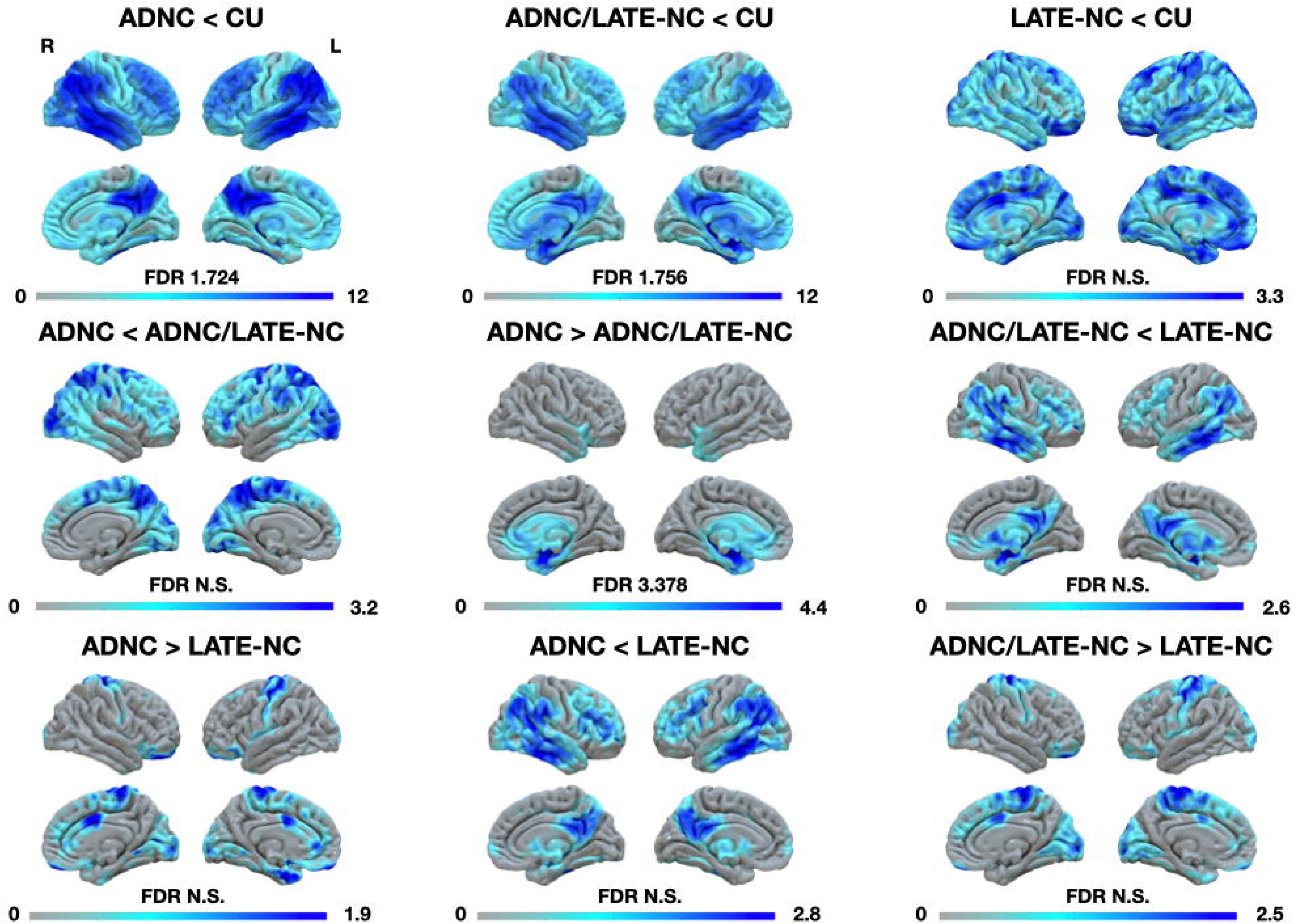
FDG-PET findings between ADNC, AD-NC/LATE-NC, LATE-NC, and CU controls. The “less than” sign reflects less metabolism in a given group relative to the other, and vice-versa. ADNC = Alzheimer’s disease neuropathological changes; LATE-NC = Limbic-predominant age-associated TDP-43 encephalopathy neuropathological changes; CU = Cognitively unimpaired; FDR = False discovery rate; N.S. = Non-significant; FDG-PET = Fluorodeoxyglucose-positron emission tomography.

### LANS likelihood differences

Table 2 indicates the distribution of ADNC, LATE-NC and ADNC/LATE-NC patients across stand-alone LANS features and LANS likelihoods for the Mayo and ADNI cohorts. These data for all pathological diagnoses are displayed in Supplementary Table 3. The assessment of differences in the distribution of LANS likelihood only applied to patients for whom all LANS features were available.

**Table 2.** LANS features and likelihoods across pathological diagnoses for the Mayo and ADNI cohorts.

| Standard and advanced LANS features (meets feature/total) |  |  |  |  |  |  |
| --- | --- | --- | --- | --- | --- | --- |
| Mayo cohort | Age $\geq 75$ | Mild syndrome | Hippocampal atrophy | Limbic hypometabolism | Absence of neocortical hypometabolism | Low neocortical tau likelihood |
| ADNC | 36/75 | 58/75 | 25/63 | 9/53 | 5/53 | 7/60 |
| ADNC/LATE-NC | 53/81 | 66/81 | 38/54 | 15/33 | 7/33 | 1/49 |
| LATE-NC | 8/9 | 8/9 | 3/4 | 3/4 | 1/4 | 5/7 |
| LANS likelihoods (only including patients with all features available) |  |  |  |  |  |  |
| Mayo cohort |  |  |  |  |  |  |
| Diagnosis | Low |  | Moderate | High |  | Highest |
| ADNC | 31 |  | 17 | 1 |  | 0 |
| ADNC/LATE-NC | 12 |  | 8 | 13 |  | 0 |
| LATE-NC | 0 |  | 0 | 2 |  | 2 |
| Standard and advanced LANS features |  |  |  |  |  |  |
| ADNI Cohort | Age $\geq 75$ | Mild syndrome | Hippocampal atrophy | Limbic hypometabolis m | Absence of neocortical hypometabolism | Low neocortical tau likelihood |
| ADNC | 15/26 | 20/26 | 11/26 | 1/18 | 2/18 | 1/22 |
| ADNC/LATE-NC | 12/19 | 12/19 | 15/19 | 4/13 | 1/13 | 0/17 |
| LATE-NC | 4/8 | 8/8 | 5/8 | 5/6 | 1/6 | 8/8 |
| LANS likelihoods (only including patients with all features available) |  |  |  |  |  |  |
| Diagnosis | Low |  | Moderate | High |  | Highest |
| ADNC | 11 |  | 7 | 0 |  | 0 |
| ADNC/LATE-NC | 8 |  | 3 | 1 |  | 0 |
| LATE-NC | 2 |  | 0 | 1 |  | 3 |

LANS = Limbic-predominant amnesic neurodegenerative syndrome; ADNC = Alzheimer's disease neuropathological changes; LATE-NC = Limbic-predominant age-related TDP-43 encephalopathy neuropathological changes.

In the Mayo cohort, all LATE-NC (4/4) patients had a highest/high likelihood. 31/49 (63%), 17/49 (35%) and 1/49 (2%) of ADNC patients had low, moderate and highest/high likelihoods, respectively. 12/33 (36%), 8/33 (24%) and 13/33 (39%) of ADNC/LATE-NC patients had low, moderate and highest/high likelihoods, respectively. Assessments of the distribution of pathological diagnoses within likelihood categories revealed that there were proportionally more ADNC/LATE-NC than ADNC patients within the highest/high likelihood category. There were more ADNC and ADNC/LATE-NC patients than LATE-NC patients within the moderate likelihood category. There were more ADNC and ADNC/LATE-NC patients than LATE-NC patients and more ADNC patients than ADNC/LATE-NC patients within the low likelihood category. Assessment of the distribution of likelihood categories within pathological diagnoses revealed that there were more ADNC patients with low and moderate likelihoods compared to highest/high likelihoods. There was no other difference.

In the ADNI cohort, 4/6 (67%) and 2/6 (33%) of LATE-NC cases had highest/high and low likelihoods, respectively. 11/18 (61%) and 7/18 (39%) of ADNC cases had low and moderate likelihoods, respectively. 8/12 (67%), 3/12 (25%) and 1/12 (8%) of ADNC/LATE-NC patients had low, moderate and highest/high likelihoods, respectively. Assessments of the distribution of pathological diagnoses within likelihood categories only revealed a trend towards more ADNC patients than LATE-NC patients within the moderate likelihood category. Assessment of the distribution of likelihood categories within pathological diagnoses revealed that there were more ADNC patients with a low likelihood compared to highest/high likelihoods. There was no other difference.

Assessment of longitudinal trajectories of CDR-SB according to LANS likelihood and FDG-PET findings comparing LANS likelihoods to CUs are displayed in Figure 3. Results from mixed linear modelling between longitudinal CDR-SB and LANS likelihoods are in Supplementary Table 4. This analysis showed that all groups had equivalent CDR-SB scores at baseline. LANS likelihoods differed in their CDR-SB trajectory over time in that patients with a high likelihood had a slower increase of CDR-SB score than those with a low likelihood. There were trends towards significance for a slower increase of CDR-SB scores in patients with a high likelihood compared to those with moderate and low likelihoods.

**Figure 3.**
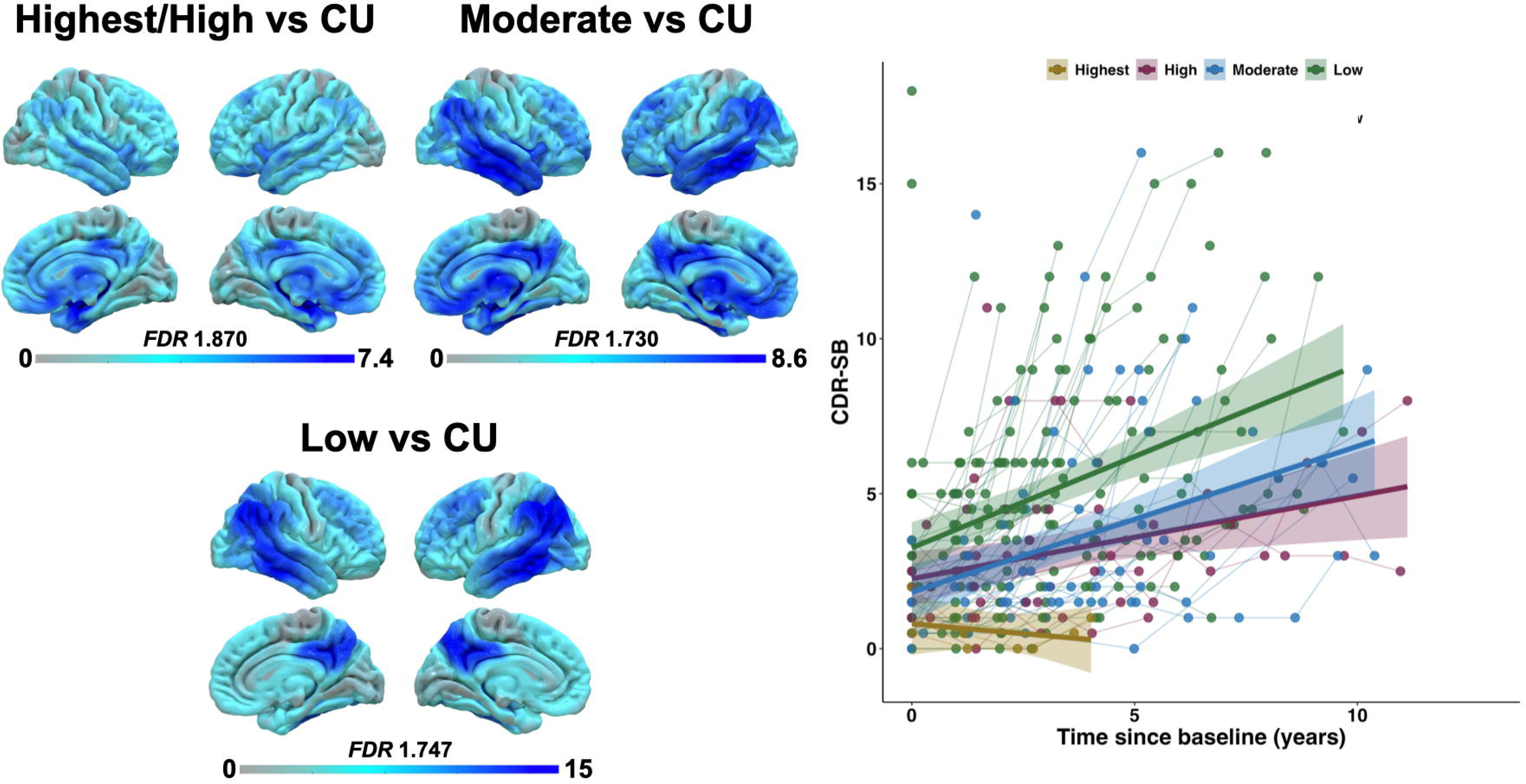
FDG-PET and longitudinal CDR-SB findings in across LANS likelihoods. FDG-PET findings comparing LANS likelihoods to CUs are displayed on the left. Longitudinal CDR-SB trajectories at the individual- and group-level are displayed on the right. CU = Cognitively unimpaired; FDR = False discovery rate; CDR-SB = Clinical dementia Rating scale – Sum of Boxes.

FDG-PET findings showed that patients with highest/high likelihoods had patterns of degeneration mostly involving the temporo-limbic system and inferior frontal areas with little involvement of neocortical areas compared to CUs. In contrast, those with a low likelihood had more pronounced neocortical degeneration mostly involving lateral temporo-parietal areas with little involvement of the medial temporal lobe. The moderate likelihood showed a mixture of these patterns, with involvement of both medial temporo-limbic and neocortical areas.

### Binary classification of LATE-NC

A logistic model classifying patients with (LATE-NC, ADNC/LATE-NC) or without LATE-NC (ADNC) based on operationalized LANS criteria (age at examination, CDR-SB score, hippocampal volume, IMT ratio, FDG-PET meta-ROI, tau positivity) achieved a balanced accuracy of 74.6% in the Mayo cohort. The sensitivity and specificity of the model were 78.6% and 70.6%, respectively. There were 12/15 (80%) true positives (i.e., correctly classified with LATE-NC) and 11/16 (68.75%) true negatives (i.e., correctly classified without LATE-NC). Out-of-sample predictions in the ADNI cohort using the model fitted in the Mayo cohort achieved a balanced accuracy of 73.3% in the ADNI cohort. The sensitivity and specificity of the model were 60.9% and 85.7%, respectively. There were 9/15 (60%) true positives (i.e., correctly classified with LATE-NC) and 14/15 (93.33%) true negatives (i.e., correctly classified without LATE-NC).

### ADNC/LATE-NC heterogeneity

We conducted an exploratory analysis to lay the foundation for future work focused on the clinical heterogeneity of ADNC/LATE-NC patients on the premise that some have LATE-NC as a primary pathology and ADNC as a co-pathology and vice-versa. We divided ADNC/LATE-NC patients based on their LANS likelihood into highest/high (*n* = 13), moderate (*n* = 8), and low (*n* = 12) likelihoods. This was only done in the Mayo cohort given the relatively low heterogeneity of LANS likelihoods in ADNC/LATE-NC patients from the ADNI cohort. Results from these analyses are visually depicted in Figure 4.

**Figure 4.**
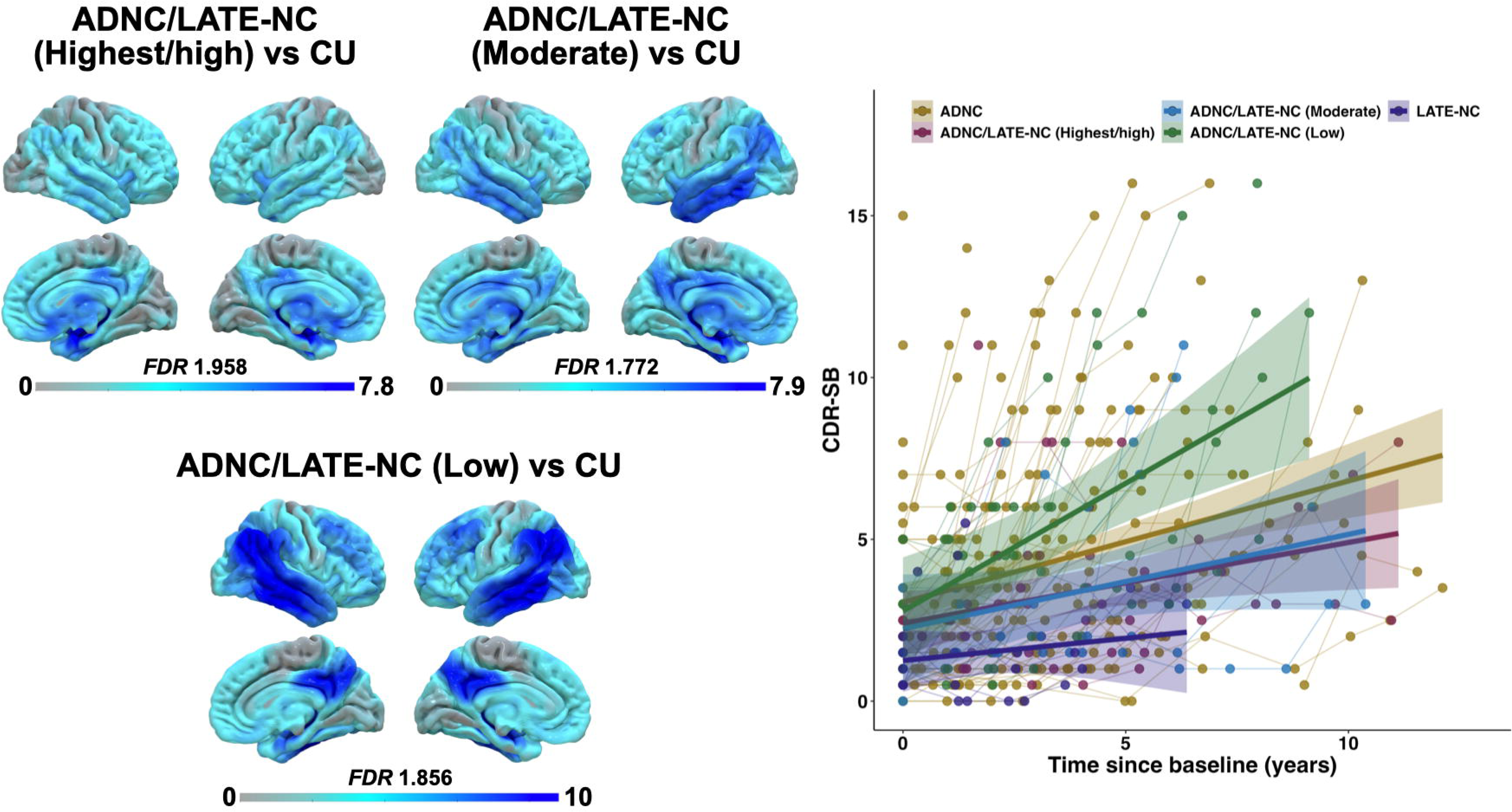
FDG-PET and longitudinal CDR-SB findings in ADNC/LATE-NC according to LANS likelihoods relative to other groups. FDG-PET findings comparing ADNC/LATE-NC to CUs are displayed on the left. Longitudinal CDR-SB trajectories at the individual- and group-level are displayed on the right. ADNC = Alzheimer’s disease neuropathological changes; LATE-NC = Limbic-predominant age-associated TDP-43 encephalopathy pathological changes; CU = Cognitively unimpaired; FDR = False discovery rate; CDR-SB = Clinical dementia Rating scale – Sum of Boxes.

Results from the mixed linear model assessing the longitudinal trajectory of CDR-SB scores according to time since baseline across groups (ADNC, ADNC/LATE-NC highest/high, ADNC/LATE-NC moderate, ADNC/LATE-NC low, LATE-NC) are in Supplementary Table 5. This analysis showed that all groups had statistically equivalent CDR-SB scores at baseline. Groups differed in their CDR-SB trajectory over time in that ADNC/LATE-NC patients with a low likelihood had a steeper increase in CDR-SB scores compared to all other groups.

FDG-PET findings showed that ADNC/LATE-NC patients with highest/high likelihoods had patterns of degeneration mostly involving the temporo-limbic system and inferior frontal areas with little involvement of neocortical areas compared to CUs. In contrast, ADNC/LATE-NC patients with a low likelihood had more pronounced neocortical degeneration mostly involving lateral temporo-parietal areas with lesser involvement of the medial temporal lobe. ADNC/LATE-NC patients with a moderate likelihood showed an intermediate pattern, with involvement of both medial temporo-limbic and neocortical areas.

## Discussion

We developed a set of clinical criteria to identify individuals with a predominant and progressive amnestic syndrome driven by degeneration of the temporo-limbic system. We termed this clinical entity “limbic-predominant amnestic neurodegenerative syndrome” or LANS to emphasize that it does not necessarily have a strict mapping to LATE-NC even though they share limbic predominance. It is also possible that there will be cases with LATE-NC that do not meet LANS criteria (e.g., patients with LBD combined with LATE-NC may meet clinical criteria for dementia with Lewy bodies and not LANS), or vice-versa. This is a degenerative neurologic syndrome with specific functional anatomic features. This neurologic condition is characterized by progressive limbic system degeneration and associated clinical signs and symptoms requiring comprehensive evaluation including history, physical, neuropsychological assessment, imaging, and fluid biomarkers. Various combinations of neuropathologies like LATE-NC, ADNC, and AGD have been observed in the setting of this neurologic syndrome. We operationalized and validated the LANS criteria by tying limbic degeneration to an older age at evaluation, mild clinical syndrome, disproportionate hippocampal atrophy according to clinical severity, limbic hypometabolism, absence of neocortical degenerative disease pattern, and low likelihood of neocortical tau pathology. The LANS criteria effectively categorized LATE-NC, ADNC/LATE-NC and ADNC patients from two clinicopathological cohorts (Mayo and ADNI). Most patients with highest or high LANS likelihoods had post-mortem evidence of LATE-NC, most patients with moderate or low LANS likelihoods had evidence of ADNC, and those with evidence of ADNC/LATE-NC patients had a wider distribution of LANS likelihoods. Assessing clinical and imaging features across LANS likelihood regardless of underlying pathology showed that patients with higher likelihoods had a slower clinical course and patterns of temporo-limbic degeneration with little involvement of neocortical areas, while those with lower likelihoods showed the opposite pattern. Stratifying ADNC/LATE-NC patients according to their LANS likelihoods revealed a high degree of heterogeneity, where those with higher likelihoods had a slower clinical course and those with lower likelihood had the worse clinical prognosis compared to all other groups. The implementation of LANS criteria has critical implications for clinical and therapeutic endeavors as well as for a deeper understanding of the etiologic landscape of predominant and progressive amnestic syndromes.

Our findings are in large agreement with previous clinicopathological studies showing associations between LATE-NC and a memory-dominant profile characterized by an indolent clinical course(6,16,17,19). They also echo studies that have found a steeper rate of cognitive decline in patients with pathological evidence for both LATE-NC and ADNC compared to those with either LATE-NC or ADNC alone(16,19). In fact, patients with LATE-NC had a mild level of cognitive impairment at presentation with relatively slow progression of impairment over time. Interestingly, patients with ADNC/LATE-NC with a high LANS likelihoods showed a relatively similar profile in terms of clinical progression, which aligns with our hypothesis that these patients have LATE-NC as the primary driver of their clinical symptoms with ADNC as a secondary pathology. This is further supported by the finding of a more rapid clinical decline in ADNC/LATE-NC patients with a low LANS likelihood, which are hypothesized to have ADNC as the primary driver of their clinical symptoms with LATE-NC as a secondary pathology. These cases decline faster because they have limbic functional neurodegeneration combined with prominent neocortical functional neurodegeneration.

FDG-PET and MRI imaging results are also indicative of a selective degeneration of the limbic system including strong involvement of the hippocampus associated with LATE-NC, above and beyond the contribution of ADNC. In fact, ADNC was significantly associated with degeneration of the lateral temporal lobe and posterior neocortical areas rather than limbic system. This aligns well with previous studies assessing imaging features of LATE-NC and/or ADNC(24,25,28–31).

The proposed LANS criteria have several clinical implications. The primary implication of these criteria is to accurately diagnose patients for whom the cause of progressive and predominant amnestic symptoms is linked to the degeneration of the limbic system with a high likelihood of LATE-NC as a symptom-driving pathology. It is noteworthy that, while the assessment of all LANS features (i.e., clinical, imaging, biomarker) is optimal to increase the diagnostic confidence of LANS, the unavailability of some features does not preclude rendering a LANS diagnosis. For instance, a routine neurological workup including a neurological assessment and an MRI study allows for the assessment of all the core and standard features. It is the clinician’s responsibility to gauge their level of confidence in a LANS diagnosis depending on available information, as is the case for any neurological condition (see Box 2 for an example of the clinical application of the LANS criteria before and after advanced features were evaluated).

The accurate diagnosis of LANS has the potential to improve prognostication and counseling of patients about the nature of their symptoms. Current evidence indicates that the symptomatology primarily involves memory for recent events while neocortical functions (e.g., executive functioning and visuospatial reasoning) are expected to remain relatively preserved throughout the disease course. An accurate diagnosis is also relevant for prognostication, as LANS has a likelihood of being associated with LATE-NC, which is associated with a relatively slow and milder clinical course compared to canonical Alzheimer’s-type dementia per our results and other studies(6,16,17,19).

To put this work in another clinical context, recent work on terminology would characterize LANS as a subtype of amnestic MCI or amnestic dementia(34). The addition of preservation of neocortical functions, older age at symptom onset, mild clinical severity and semantic memory impairment would make these amnestic syndromes more likely to be LANS and the additional imaging would localize the syndromes to the limbic system. These refinements would add greater specificity to the diagnosis for clinicians.

The LANS criteria have critical therapeutic implications. This is especially true in this era of emerging anti-amyloid monoclonal antibody therapies(32,70–72). As these therapies make their way into clinical practice, the LANS criteria can be of relevance for identifying patients with a high likelihood of LATE-NC as the primary cause of their symptoms and guide clinical decision making and counselling regarding potential therapeutic avenues. To illustrate this point, we highlight an example of the prospective application of the LANS criteria on a patient seen in clinical practice in the context of determining eligibility for an anti-amyloid monoclonal antibody therapy in Box 2. It is important to mention that positive Alzheimer’s disease biomarkers do not rule out a diagnosis of LANS, as a progressive and predominant amnestic syndrome caused by LATE-NC can be observed in concomitance with incidental amyloidosis or ADNC(28,29). The use of the LANS likelihoods can help determining whether the amnestic syndrome is most likely driven by primary LATE-NC with secondary ADNC or vice-versa, as demonstrated by our analysis deciphering the heterogeneity of patients with ADNC/LATE-NC. Future studies will be important to determine how patients with positive Alzheimer’s disease biomarkers respond to disease-modifying therapies according to their LANS likelihood. Finally, the LANS criteria are a major advancement in the overarching goal of tying an *in vivo* neurologic syndrome to underlying LATE-NC. The other critical component needed to achieve this is the development of *in vivo* diagnostic biomarkers of TDP-43 specific for LATE-NC. PET, CSF and biofluid biomarkers of TDP-43 are hopefully on the horizon(6,73,74). This would allow for defining the clinical entity of LATE as a high-likelihood LANS syndrome with biomarker evidence of LATE-NC. Even if a novel TDP-43 biomarker is not specific for LATE-NC (e.g., it may also be positive in other pathologic entities associated with TDP-43), it will still be highly specific for LATE-NC in the setting of a LANS neurologic syndrome. Defining such a clinicopathological entity will be important for the design of clinical trials aimed at LATE-NC in terms of defining eligibility criteria and outcomes measures. Prior to a LATE-NC specific biomarker, a high-likelihood LANS presentation will allow for enrichment of clinical trial populations with LATE-NC.

It is important to reiterate that while LANS is highly associated with LATE-NC and we provide an initial validation of syndrome, it can be associated with other pathologic entities that selectively target the limbic system. Supplemental analyses provide some evidence that pathologies associated with a predominant amnestic syndrome other than LATE-NC may, in rare instances, have a high LANS likelihood (e.g., Lewy Body Disease, AGD, advanced PART). One example that can be a potential source of clinical conundrums is the limbic variant Alzheimer’s disease, which localizes to the limbic system and therefore qualifies for LANS. In this scenario, the advanced LANS criteria in combination with visual assessment of tau-PET can help in determining which pathology has the highest likelihood of driving clinical symptoms. A combination of a LANS diagnosis combined with a biomarker of limbic predominate tau deposition could be used to define a clinicopathologic entity of limbic Alzheimer’s disease.

Notably, there were slightly lower LANS likelihoods in ADNC/LATE-NC patients in the ADNI cohort relative to the Mayo cohort. This is likely due to discrepancy in recruitment and sampling strategies. The Mayo cohort draws from clinical practice in a tertiary behavioral neurology clinic (ADRC) and randomly selected individuals living in Olmsted County, MN, USA (MCSA)(75). It is thus designed to reflect a combination of the clinicopathologic variability encountered in Alzheimer’s-oriented clinical context, and also with more representative settings. On the other hand, ADNI was designed as a research cohort aimed at the clinical and biological characterization of individuals on the clinicopathologic spectrum of Alzheimer’s disease(76,77). It is not meant to reflect the heterogeneity encountered in clinical practice. These fundamental differences may explain the relatively younger age of LATE-NC patients, the lower frequency of ADNC/LATE-NC with a high LANS likelihood, and the lower accuracy in the binary classification of LATE-NC in the ADNI cohort. We believe the performance of the LANS criteria in the Mayo cohort is a more accurate reflection of how the LANS criteria are expected to perform in clinical settings. It is also important to mention that we undertook several steps to avoid overfitting the LANS criteria validation in the Mayo cohort. This includes the use of externally validated thresholds for most imaging and tau biomarkers (e.g., CSF, ptau181, molecular PET imaging) and deriving optimal thresholds for hippocampal atrophy and the IMT ratio in the ADNI cohort rather than Mayo.

There are some limitations to this work. An evident caveat is the low number of patients with LATE-NC without ADNC. The lack of statistical differences between these patients and other groups in terms of clinical and imaging features could be attributable to the small sample size rather than a genuine absence of differences. However, most patients with LATE-NC had highest and high LANS likelihoods, suggesting that our proposed criteria effectively identify these individuals. Although the proposed LANS criteria include impaired semantic memory, we did not incorporate this feature in the operationalized criteria given the less robust and variably defined evidence of such impairment. Efforts are underway to better characterize the nature and extent of semantic impairment in patients with LATE-NC and develop cognitive tests sensitive to such impairment. This should not, however, prevent clinicians from considering this feature in their clinical decision making when LANS is in the differential, as highlighted in the case example in Box 2. This study is retrospective in nature. The implementation of the LANS criteria in clinical settings and prospective studies are needed to further validate and refine this set of criteria.

In conclusion, we developed and validated a set of criteria for LANS designed to be used in clinical practice to identify individuals with a predominant amnestic syndrome driven by the degeneration of the temporo-limbic system with a high likelihood of underlying LATE-NC. This has important clinical implications including differential diagnosis with other common causes of memory impairment such as ADNC, counselling patients about the nature and course of their symptoms, providing appropriate treatments, referral to research programs, and the development of therapeutic efforts aimed at LATE-NC. Several steps lay ahead to improve the definition of LANS including the conduction of prospective studies and the development of clinical tools that are sensitive and specific to its cognitive features. Finally, the development of *in vivo* diagnostic markers of TDP-43 pathology is needed to embed LANS into a clinicopathological entity driven by LATE-NC, i.e., LATE.

## Data Availability

All data produced in the present study are available upon reasonable request to the authors.

https://www.mayo.edu/research/centers-programs/alzheimers-disease-research-center/data-requests

https://adni.loni.usc.edu/

https://www.nitrc.org/projects/mcalt/

## Acknowledgements

We wish to thank our patients and their caregivers for their dedicated participation in our research program. We would also wish to express our gratitude to all healthcare providers and research professionals who were involved in this study and patient care.

## Supplementary material

Supplementary material is available at *Brain* online.

## Box 1

### Limbic-predominant amnestic neurodegenerative syndrome (LANS)

#### Clinical criteria

##### Core clinical features

Must present with a slow, amnestic, predominant neurodegenerative syndrome (insidious onset with gradual progression over two or more years) without another condition that better accounts for the clinical deficits.

###### Standard supportive features

1. Older age at evaluation (generally ≥75-year-old);
2. Mild clinical syndrome with largely preserved neocortical-predominant functions;
3. Hippocampal atrophy out of proportion to syndrome severity;
4. Impaired semantic memory in the setting of a mild syndrome.

###### Advanced supportive features

1. Limbic hypometabolism and absence of neocortical degenerative pattern on FDG-PET imaging;
2. Low likelihood of significant neocortical tau pathology.

###### Degree of certainty

1. Low-likelihood: Meets core features and ≤2 standard features;
2. Moderate-likelihood: Meets core features and ≥3 standard features, or meets core features and ≥2 standard and 1 advanced features;
3. High-likelihood: Meets core features, ≥3 standard features, and 1 advanced feature, or meets core features, ≥2 standard features, and 2 advanced features
4. Highest-likelihood: Meets all core, standard and advanced features.

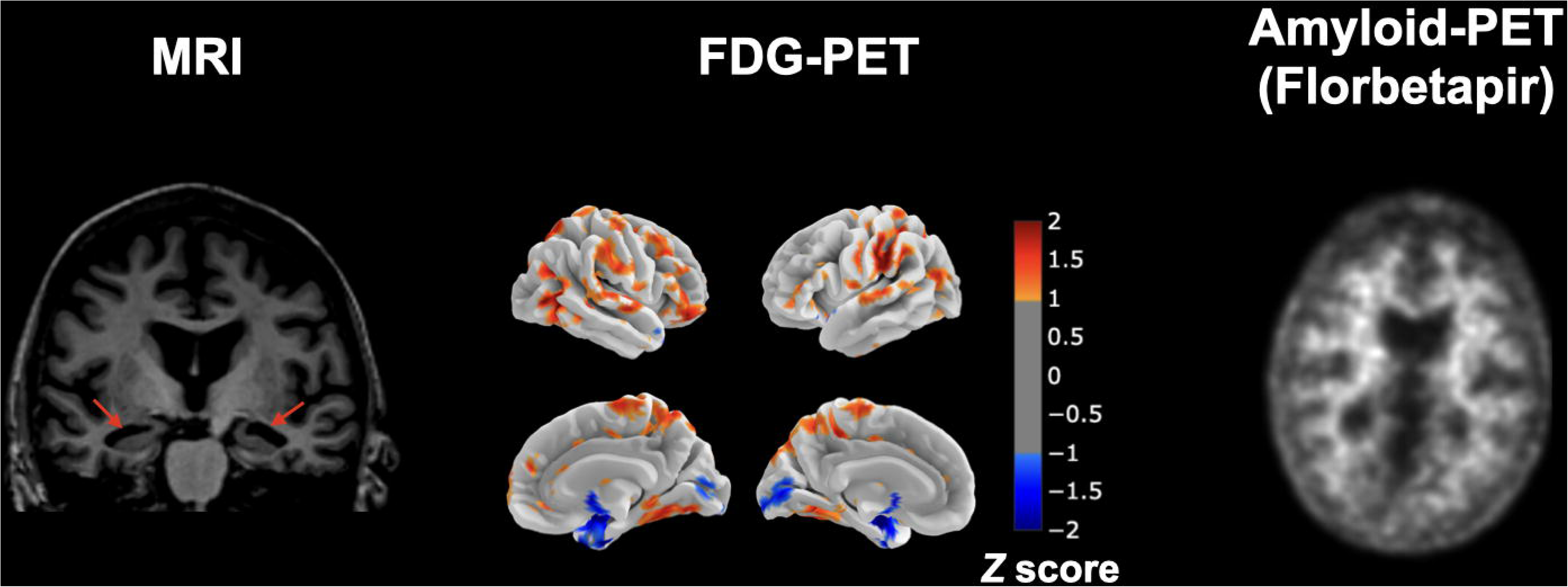

## Box 2

### Case example of a prospective use of the LANS criteria in clinical settings.

This patient was seen at Mayo Clinic Rochester to determine eligibility for an anti-amyloid monoclonal antibody therapy, and the LANS criteria were internally defined at the time of evaluation. The criteria were operationalized using clinical judgment of available data including visual reads of neuroimaging as is the current standard in clinical practice. This patient was a female over the age of 75 with a history of memory problems for several years. She lived alone, managed her instrumental activities of daily living independently and was still working part-time. Neuropsychological testing revealed moderate to severe impairment on measures of delayed recall and fragility of salient semantic knowledge (i.e., she could not recall details about the events occurring on 9/11/2001; she named another building than the World Trade Center and did not know how many buildings were hit). Performance was also mildly low on a task of object naming. The remainder of the assessment (i.e., global cognition, executive functions, visuospatial processing and language) was within or above expectations from a normative standpoint. The profile was consistent with single-domain amnestic mild cognitive impairment. MRI revealed disproportionate bilateral hippocampal atrophy, and she was assigned a moderate likelihood LANS diagnosis. Subsequently, a brain FDG-PET revealed prominent temporo-limbic and milder inferior frontal hypometabolism in the absence of a neocortical degenerative pattern by visual read. Amyloid-PET was read as negative. She thus met all core, standard and advanced criteria for LANS, corresponding to the highest likelihood. She did not meet criteria for an anti-amyloid monoclonal antibody therapy.

## References

1. Nelson PT, Dickson DW, Trojanowski JQ, Jack CR, Boyle PA, Arfanakis K, et al. Limbic-predominant age-related TDP-43 encephalopathy (LATE): Consensus working group report. Brain. 2019;142(6):1503–27.

2. Josephs KA, Ahmed Z, Katsuse O, Parisi JF, Boeve BF, Knopman DS, et al. Neuropathologic features of frontotemporal lobar degeneration with ubiquitin-positive inclusions with progranulin gene (PGRN) mutations. J Neuropathol Exp Neurol. 2007;66(2):142–51.

3. Josephs KA, Murray ME, Tosakulwong N, Weigand SD, Serie AM, Perkerson RB, et al. Pathological, imaging and genetic characteristics support the existence of distinct TDP-43 types in non-FTLD brains. Acta Neuropathol. 2019;137:227–38.

4. Josephs KA, Murray ME, Whitwell JL, Parisi JE, Petrucelli L, Jack CR, et al. Staging TDP-43 pathology in Alzheimer’s disease. Acta Neuropathol. 2014;127:441–50.

5. Gauthreaux KM, Teylan MA, Katsumata Y, Mock C, Culhane JE, Chen Y-C, et al. Limbic-predominant age-related TDP-43 encephalopathy: medical and pathologic factors associated with comorbid hippocampal sclerosis. Neurology. 2022;98(14):e1422–33.

6. Duong MT, Wolk DA. Limbic-predominant age-related TDP-43 encephalopathy: LATE-breaking updates in clinicopathologic features and biomarkers. Curr Neurol Neurosci Rep. 2022;22(11):689–98.

7. Dickson DW, Davies P, Bevona C, Van Hoeven KH, Factor SM, Grober E, et al. Hippocampal sclerosis: a common pathological feature of dementia in very old (≥ 80 years of age) humans. Acta Neuropathol. 1994;88:212–21.

8. Young AL, Vogel JW, Robinson JL, McMillan CT, Ossenkoppele R, Wolk DA, et al. Data-driven neuropathological staging and subtyping of TDP-43 proteinopathies. Brain. 2023;146(7):2975–88.

9. Josephs KA, Murray ME, Whitwell JL, Tosakulwong N, Weigand SD, Petrucelli L, et al. Updated TDP-43 in Alzheimer’s disease staging scheme. Acta Neuropathol. 2016;131:571–85.

10. Coyle-Gilchrist ITS, Dick KM, Patterson K, Rodríquez PV, Wehmann E, Wilcox A, et al. Prevalence, characteristics, and survival of frontotemporal lobar degeneration syndromes. Neurology. 2016;86(18):1736–43.

11. Nelson PT, Brayne C, Flanagan ME, Abner EL, Agrawal S, Attems J, et al. Frequency of LATE neuropathologic change across the spectrum of Alzheimer’s disease neuropathology: Combined data from 13 community-based or population-based autopsy cohorts. Acta Neuropathol. 2022;144(1):27–44.

12. Robinson JL, Lee EB, Xie SX, Rennert L, Suh E, Bredenberg C, et al. Neurodegenerative disease concomitant proteinopathies are prevalent, age-related and APOE4-associated. Brain. 2018;141(7):2181–93.

13. Wennberg AM, Tosakulwong N, Lesnick TG, Murray ME, Whitwell JL, Liesinger AM, et al. Association of apolipoprotein E ε4 with transactive response DNA-binding protein 43. JAMA Neurol. 2018;75(11):1347–54.

14. Yang H-S, Yu L, White CC, Chibnik LB, Chhatwal JP, Sperling RA, et al. Evaluation of TDP-43 proteinopathy and hippocampal sclerosis in relation to APOE ε4 haplotype status: a community-based cohort study. Lancet Neurol. 2018;17(9):773–81.

15. Josephs KA, Whitwell JL, Weigand SD, Murray ME, Tosakulwong N, Liesinger AM, et al. TDP-43 is a key player in the clinical features associated with Alzheimer’s disease. Acta Neuropathol. 2014;127:811–24.

16. Gauthreaux K, Mock C, Teylan MA, Culhane JE, Chen Y-C, Chan KCG, et al. Symptomatic profile and cognitive performance in autopsy-confirmed limbic-predominant age-related TDP-43 encephalopathy with comorbid Alzheimer disease. J Neuropathol Exp Neurol. 2022;81(12):975–87.

17. Kapasi A, Yu L, Boyle PA, Barnes LL, Bennett DA, Schneider JA. Limbic-predominant age-related TDP-43 encephalopathy, ADNC pathology, and cognitive decline in aging. Neurology. 2020;95(14):e1951–62.

18. Brenowitz WD, Monsell SE, Schmitt FA, Kukull WA, Nelson PT. Hippocampal sclerosis of aging is a key Alzheimer’s disease mimic: clinical-pathologic correlations and comparisons with both Alzheimer’s disease and non-tauopathic frontotemporal lobar degeneration. J Alzheimer’s Dis. 2014;39(3):691–702.

19. Lopez OL, Kofler J, Chang Y, Berman SB, Becker JT, Sweet RA, et al. Hippocampal sclerosis, TDPL43, and the duration of the symptoms of dementia of AD patients. Ann Clin Transl Neurol. 2020;7(9):1546–56.

20. Katsumata Y, Abner EL, Karanth S, Teylan MA, Mock CN, Cykowski MD, et al. Distinct clinicopathologic clusters of persons with TDP-43 proteinopathy. Acta Neuropathol. 2020;140:659–74.

21. Karanth S, Nelson PT, Katsumata Y, Kryscio RJ, Schmitt FA, Fardo DW, et al. Prevalence and clinical phenotype of quadruple misfolded proteins in older adults. JAMA Neurol. 2020;77(10):1299–307.

22. Buciuc M, Tosakulwong N, Machulda MM, Whitwell JL, Weigand SD, Murray ME, et al. TAR DNA-binding protein 43 is associated with rate of memory, functional and global cognitive decline in the decade prior to death. J Alzheimer’s Dis. 2021;80(2):683–93.

23. Wilson RS, Yu L, Trojanowski JQ, Chen E-Y, Boyle PA, Bennett DA, et al. TDP-43 pathology, cognitive decline, and dementia in old age. JAMA Neurol. 2013;70(11):1418– 24.

24. Josephs KA, Martin PR, Weigand SD, Tosakulwong N, Buciuc M, Murray ME, et al. Protein contributions to brain atrophy acceleration in Alzheimer’s disease and primary age-related tauopathy. Brain. 2020;143(11):3463–76.

25. Buciuc M, Wennberg AM, Weigand SD, Murray ME, Senjem ML, Spychalla AJ, et al. Effect modifiers of TDP-43-associated hippocampal atrophy rates in patients with Alzheimer’s disease neuropathological changes. J Alzheimer’s Dis. 2020;73(4):1511–23.

26. Bejanin A, Murray ME, Martin P, Botha H, Tosakulwong N, Schwarz CG, et al. Antemortem volume loss mirrors TDP-43 staging in older adults with non-frontotemporal lobar degeneration. Brain. 2019;142(11):3621–35.

27. AmadorLOrtiz C, Lin W, Ahmed Z, Personett D, Davies P, Duara R, et al. TDPL43 immunoreactivity in hippocampal sclerosis and Alzheimer’s disease. Ann Neurol Off J Am Neurol Assoc Child Neurol Soc. 2007;61(5):435–45.

28. Botha H, Mantyh WG, Murray ME, Knopman DS, Przybelski SA, Wiste HJ, et al. FDG-PET in tau-negative amnestic dementia resembles that of autopsy-proven hippocampal sclerosis. Brain. 2018;141(4):1201–17.

29. Botha H, Mantyh WG, Graff-Radford J, Machulda MM, Przybelski SA, Wiste HJ, et al. Tau-negative amnestic dementia masquerading as Alzheimer disease dementia. Neurology. 2018;90(11):e940–6.

30. Buciuc M, Botha H, Murray ME, Schwarz CG, Senjem ML, Jones DT, et al. Utility of FDG-PET in diagnosis of Alzheimer-related TDP-43 proteinopathy. Neurology. 2020;95(1):23–34.

31. Grothe MJ, Moscoso A, SilvaLRodríguez J, Lange C, Nho K, Saykin AJ, et al. Differential diagnosis of amnestic dementia patients based on an FDGLPET signature of autopsyLconfirmed LATELNC. Alzheimer’s Dement. 2023;19(4):1234–44.

32. Ramanan VK, Armstrong MJ, Choudhury P, Coerver K, Hamilton RH, Klein BC, et al. Antiamyloid Monoclonal Antibody Therapy for Alzheimer Disease: Emerging Issues in Neurology. Neurology. 2023;

33. Jones D, Lowe V, Graff-Radford J, Botha H, Barnard L, Wiepert D, et al. A computational model of neurodegeneration in Alzheimer’s disease. Nat Commun [Internet]. 2022;13(1):1643. Available from: http://www.ncbi.nlm.nih.gov/pubmed/35347127

34. Petersen RC, Weintraub S, Sabbagh M, Karlawish J, Adler CH, Dilworth-Anderson P, et al. A New Framework for Dementia Nomenclature. JAMA Neurol. 2023;

35. McKhann GM, Knopman DS, Chertkow H, Hyman BT, Jack CR, Kawas CH, et al. The diagnosis of dementia due to Alzheimer’s disease: Recommendations from the National Institute on Aging-Alzheimer’s Association workgroups on diagnostic guidelines for Alzheimer’s disease. Alzheimer’s Dement [Internet]. 2011;7(3):263–9. Available from: 10.1016/j.jalz.2011.03.005

36. Albert MS, DeKosky ST, Dickson D, Dubois B, Feldman HH, Fox NC, et al. The diagnosis of mild cognitive impairment due to Alzheimer’s disease: Recommendations from the National Institute on Aging-Alzheimer’s Association workgroups on diagnostic guidelines for Alzheimer’s disease. Alzheimer’s Dement [Internet]. 2011;7(3):270–9. Available from: 10.1016/j.jalz.2011.03.008

37. Petersen RC, Doody R, Kurz A, Mohs RC, Morris JC, Rabins P V., et al. Current concepts in mild cognitive impairment. Arch Neurol. 2001;58(12):1985–92.

38. Mirra SS, Heyman A, McKeel D, Sumi SM, Crain BJ, Brownlee LM, et al. The Consortium to Establish a Registry for Alzheimer’s Disease (CERAD): Part II. Standardization of the neuropathologic assessment of Alzheimer’s disease. Neurology. 1991;41(4):479.

39. Hyman BT, Phelps CH, Beach TG, Bigio EH, Cairns NJ, Carrillo MC, et al. National Institute on Aging–Alzheimer’s Association guidelines for the neuropathologic assessment of Alzheimer’s disease. Alzheimer’s Dement. 2012;8(1):1–13.

40. Braak H, Alafuzoff I, Arzberger T, Kretzschmar H, Del Tredici K. Staging of Alzheimer disease-associated neurofibrillary pathology using paraffin sections and immunocytochemistry. Acta Neuropathol. 2006;112(4):389–404.

41. Crary JF, Trojanowski JQ, Schneider JA, Abisambra JF, Abner EL, Alafuzoff I, et al. Primary age-related tauopathy (PART): a common pathology associated with human aging. Acta Neuropathol. 2014;128(6):755–66.

42. Mackenzie IRA, Neumann M, Baborie A, Sampathu DM, Du Plessis D, Jaros E, et al. A harmonized classification system for FTLD-TDP pathology. Acta Neuropathol. 2011;122(1):111–3.

43. Montine TJ, Phelps CH, Beach TG, Bigio EH, Cairns NJ, Dickson DW, et al. National Institute on Aging–Alzheimer’s Association guidelines for the neuropathologic assessment of Alzheimer’s disease: a practical approach. Acta Neuropathol. 2012;123(1):1–11.

44. Dickson DW, Bergeron C, Chin SS, Duyckaerts C, Horoupian D, Ikeda K, et al. Office of Rare Diseases neuropathologic criteria for corticobasal degeneration. J Neuropathol Exp Neurol. 2002;61(11):935–46.

45. Briggs M, Allinson KSJ, Malpetti M, Spillantini MG, Rowe JB, Kaalund SS. Validation of the new pathology staging system for progressive supranuclear palsy. Acta Neuropathol. 2021;141:787–9.

46. Jellinger KA. Dementia with grains (argyrophilic grain disease). Brain Pathol. 1998;8(2):377–86.

47. Stricker NH, Stricker JL, Karstens AJ, Geske JR, Fields JA, Hassenstab J, et al. A novel computer adaptive word list memory test optimized for remote assessment: Psychometric properties and associations with neurodegenerative biomarkers in older women without dementia. Alzheimer’s Dement Diagnosis, Assess Dis Monit. 2022;14(1):e12299.

48. Jack Jr CR, Wiste HJ, Weigand SD, Knopman DS, Mielke MM, Vemuri P, et al. Different definitions of neurodegeneration produce similar amyloid/neurodegeneration biomarker group findings. Brain. 2015;138(12):3747–59.

49. Jack CR, Wiste HJ, Weigand SD, Therneau TM, Lowe VJ, Knopman DS, et al. Defining imaging biomarker cut points for brain aging and Alzheimer’s disease. Alzheimer’s Dement. 2017;13(3):205–16.

50. Landau SM, Harvey D, Madison CM, Reiman EM, Foster NL, Aisen PS, et al. Comparing predictors of conversion and decline in mild cognitive impairment. Neurology. 2010;75(3):230–8.

51. Schwarz CG, Gunter JL, Wiste HJ, Przybelski SA, Weigand SD, Ward CP, et al. A large-scale comparison of cortical thickness and volume methods for measuring Alzheimer’s disease severity. NeuroImage Clin [Internet]. 2016;11:802–12. Available from: 10.1016/j.nicl.2016.05.017

52. Lowe VJ, Lundt ES, Albertson SM, Min H-K, Fang P, Przybelski SA, et al. Tau-positron emission tomography correlates with neuropathology findings. Alzheimer’s Dement. 2019;

53. Landau SM, Mintun MA, Joshi AD, Koeppe RA, Petersen RC, Aisen PS, et al. Amyloid deposition, hypometabolism, and longitudinal cognitive decline. Ann Neurol. 2012;72(4):578–86.

54. Royse SK, Minhas DS, Lopresti BJ, Murphy A, Ward T, Koeppe RA, et al. Validation of amyloid PET positivity thresholds in centiloids: a multisite PET study approach. Alzheimers Res Ther. 2021;13(1):1–10.

55. Lowe VJ, Lundt ES, Albertson SM, Min HK, Fang P, Przybelski SA, et al. Tau-positron emission tomography correlates with neuropathology findings. Alzheimer’s Dement [Internet]. 2020;16(3):561–71. Available from: 10.1016/j.jalz.2019.09.079

56. Van Harten AC, Wiste HJ, Weigand SD, Mielke MM, Kremers WK, Eichenlaub U, et al. CSF biomarkers in Olmsted County: evidence of 2 subclasses and associations with demographics. Neurology. 2020;95(3):e256–67.

57. Bittner T, Zetterberg H, Teunissen CE, Ostlund Jr RE, Militello M, Andreasson U, et al. Technical performance of a novel, fully automated electrochemiluminescence immunoassay for the quantitation of β-amyloid (1–42) in human cerebrospinal fluid. Alzheimer’s Dement. 2016;12(5):517–26.

58. Blennow K, Shaw LM, Stomrud E, Mattsson N, Toledo JB, Buck K, et al. Predicting clinical decline and conversion to Alzheimer’s disease or dementia using novel Elecsys Aβ (1–42), pTau and tTau CSF immunoassays. Sci Rep. 2019;9(1):19024.

59. Hansson O, Seibyl J, Stomrud E, Zetterberg H, Trojanowski JQ, Bittner T, et al. CSF biomarkers of Alzheimer’s disease concord with amyloid-β PET and predict clinical progression: a study of fully automated immunoassays in BioFINDER and ADNI cohorts. Alzheimer’s Dement. 2018;14(11):1470–81.

60. Bermudez C, Graff-Radford J, Syrjanen JA, Stricker NH, Algeciras-Schimnich A, Kouri N, et al. Plasma biomarkers for prediction of Alzheimer’s disease neuropathologic change. Acta Neuropathol. 2023;1–17.

61. Mattsson N, Cullen NC, Andreasson U, Zetterberg H, Blennow K. Association between longitudinal plasma neurofilament light and neurodegeneration in patients with Alzheimer disease. JAMA Neurol. 2019;76(7):791–9.

62. Mattsson N, Andreasson U, Zetterberg H, Blennow K, Initiative ADN. Association of plasma neurofilament light with neurodegeneration in patients with Alzheimer disease. JAMA Neurol. 2017;74(5):557–66.

63. Karikari TK, Pascoal TA, Ashton NJ, Janelidze S, Benedet AL, Rodriguez JL, et al. Blood phosphorylated tau 181 as a biomarker for Alzheimer’s disease: a diagnostic performance and prediction modelling study using data from four prospective cohorts. Lancet Neurol. 2020;19(5):422–33.

64. Karikari TK, Benedet AL, Ashton NJ, Lantero Rodriguez J, Snellman A, Suárez-Calvet M, et al. Diagnostic performance and prediction of clinical progression of plasma phospho-tau181 in the Alzheimer’s Disease Neuroimaging Initiative. Mol Psychiatry. 2021;26(2):429–42.

65. O’Bryant SE, Lacritz LH, Hall J, Waring SC, Chan W, Khodr ZG, et al. Validation of the new interpretive guidelines for the clinical dementia rating scale sum of boxes score in the national Alzheimer’s coordinating center database. Arch Neurol. 2010;67(6):746–9.

66. Jack CR, Wiste HJ, Botha H, Weigand SD, Therneau TM, Knopman DS, et al. The bivariate distribution of amyloid-β and tau: Relationship with established neurocognitive clinical syndromes. Brain. 2019;142(10):3230–42.

67. Tondo G, Carli G, Santangelo R, Mattoli MV, Presotto L, Filippi M, et al. BiomarkerLbased stability in limbicLpredominant amnestic mild cognitive impairment. Eur J Neurol. 2021;28(4):1123–33.

68. Caminiti SP, De Francesco S, Tondo G, Galli A, Redolfi A, Perani D, et al. FDGLPET markers of heterogeneity and different risk of progression in amnestic MCI. Alzheimer’s Dement. 2023;

69. Jack Jr CR, Wiste HJ, Algeciras-Schimnich A, Figdore DJ, Schwarz CG, Lowe VJ, et al. Predicting amyloid PET and tau PET stages with plasma biomarkers. Brain. 2023;146(5):2029–44.

70. Knopman DS, Hershey L. Implications of the Approval of Lecanemab for Alzheimer Disease Patient Care: Incremental Step or Paradigm Shift. Neurology. 2023;

71. Van Dyck CH, Swanson CJ, Aisen P, Bateman RJ, Chen C, Gee M, et al. Lecanemab in early Alzheimer’s disease. N Engl J Med. 2023;388(1):9–21.

72. Sims JR, Zimmer JA, Evans CD, Lu M, Ardayfio P, Sparks J, et al. Donanemab in early symptomatic Alzheimer disease: the TRAILBLAZER-ALZ 2 randomized clinical trial. Jama. 2023;330(6):512–27.

73. Feneberg E, Gray E, Ansorge O, Talbot K, Turner MR. Towards a TDP-43-based biomarker for ALS and FTLD. Mol Neurobiol. 2018;55:7789–801.

74. Kapaki E, Boufidou F, Bourbouli M, Pyrgelis E-S, Constantinides VC, Anastassopoulou C, et al. Cerebrospinal fluid biomarker profile in TDP-43-related genetic frontotemporal dementia. J Pers Med. 2022;12(10):1747.

75. Roberts RO, Geda YE, Knopman DS, Cha RH, Pankratz VS, Boeve BF, et al. The Mayo Clinic Study of Aging: design and sampling, participation, baseline measures and sample characteristics. Neuroepidemiology. 2008;30(1):58–69.

76. Petersen RC, Aisen PS, Beckett LA, Donohue MC, Gamst AC, Harvey DJ, et al. Alzheimer’s disease neuroimaging initiative (ADNI): clinical characterization. Neurology. 2010;74(3):201–9.

77. Mueller SG, Weiner MW, Thal LJ, Petersen RC, Jack CR, Jagust W, et al. Ways toward an early diagnosis in Alzheimer’s disease: the Alzheimer’s Disease Neuroimaging Initiative (ADNI). Alzheimer’s Dement. 2005;1(1):55–66.

