## Supplementary Materials for "A limbic-predominant amnestic neurodegenerative syndrome associated with TDP-43 pathology"

**Imaging acquisition protocols**

MRI scans acquired at Mayo Clinic were performed with a General Electronics (GE) (1.5T or 3T) scanner using a magnetization prepared rapid gradient echo sequence. Acquisition parameters included the following: repetition time = 2300 milliseconds, echo time = 3 milliseconds, T1 = 900 milliseconds, flip angle = 8^o^, field of view = 26 centimeters, 256 X 256 in-plane matrix with a phase field of 0.94, slice thickness = 1.2 millimeters. MRI scans from ADNI were acquired using 3T scanners using a magnetization prepared rapid gradient echo sequence with 1 millimeter isotropic voxel size and a time of repetition of 2300 milliseconds (more detail can be found on <https://adni.loni.usc.edu/wp-content/uploads/2017/07/ADNI3-MRI-protocols.pdf>).

FDG-PET scans acquired at Mayo Clinic were performed using a PET/CT scanner (GE healthcare or Siemens) following a 30-minute uptake period while waiting in a dim lit root. The scanning session lasted 8 minutes divided into four 2-minutes dynamic frames following a low-dose CT transmission scan. FDG-PET scans from ADNI were acquired during a 30-minutes dynamic emission scan divided in six 5-minutes frames following a 30-60 minutes uptake period following the injection of [^18^F]FDG-PET (more detail can be found on <https://adni.loni.usc.edu/methods/pet-analysis-method/pet-analysis/>).

Amyloid-PET scans acquired at Mayo Clinic were performed using the Pittsburgh compound B (PiB) ligand whereas those acquired in the context of ADNI were performed using either the Florbetapir (Amyvid) or Florbetaben (Neuraceq) ligands. Tau-PET images were using and Flortaucipir for both the Mayo Clinic and ADNI cohorts. Acquisition protocols and processing for the Mayo cohort are described in separate publications^1,2^, and acquisition protocols for the ADNI cohort can be found <https://adni.loni.usc.edu/methods/pet-analysis-method/pet-analysis/>.

**Neuropathological assessment protocols**

Regarding the Mayo cohort, immunochemistry was done using a battery of antibodies for a-synuclein (rabbit polyclonal; NACP, Mayo Clinic antibody, 1:3000 with 95% formic acid pretreatment and DAKO EnVision reagents; Carpinteria, CA), phosphorylated TDP-43 antibody (pS409/410, 1:5000 mouse monoclonal, Cosmo Bio Co., LFTD), Aβ (6F/3D, 1:250, human AB8-17, DAKO, Carpinteria, CA), P-tau (CP13, 1:1000, IgG1 to phosphor-serine 202, Long Island, NY), 4R-tau (RD4, 1:5000) and 3R-tau (RD3, 1:5000) (Millipore, Temecula, CA). Senile plaques and neurofibrillary tangles were assessed with thioflavin S fluorescent microscopy and Bielschowsky and Gallyas silver stains, respectively. Neuropathological assessments from the ADNI cohort were performed according to established protocols and disease staging was performed according to established criteria. These procedures are comprehensively described in separate publications^3–5^ and on the following page: <https://adni.loni.usc.edu/methods/neuropath-methods/>.

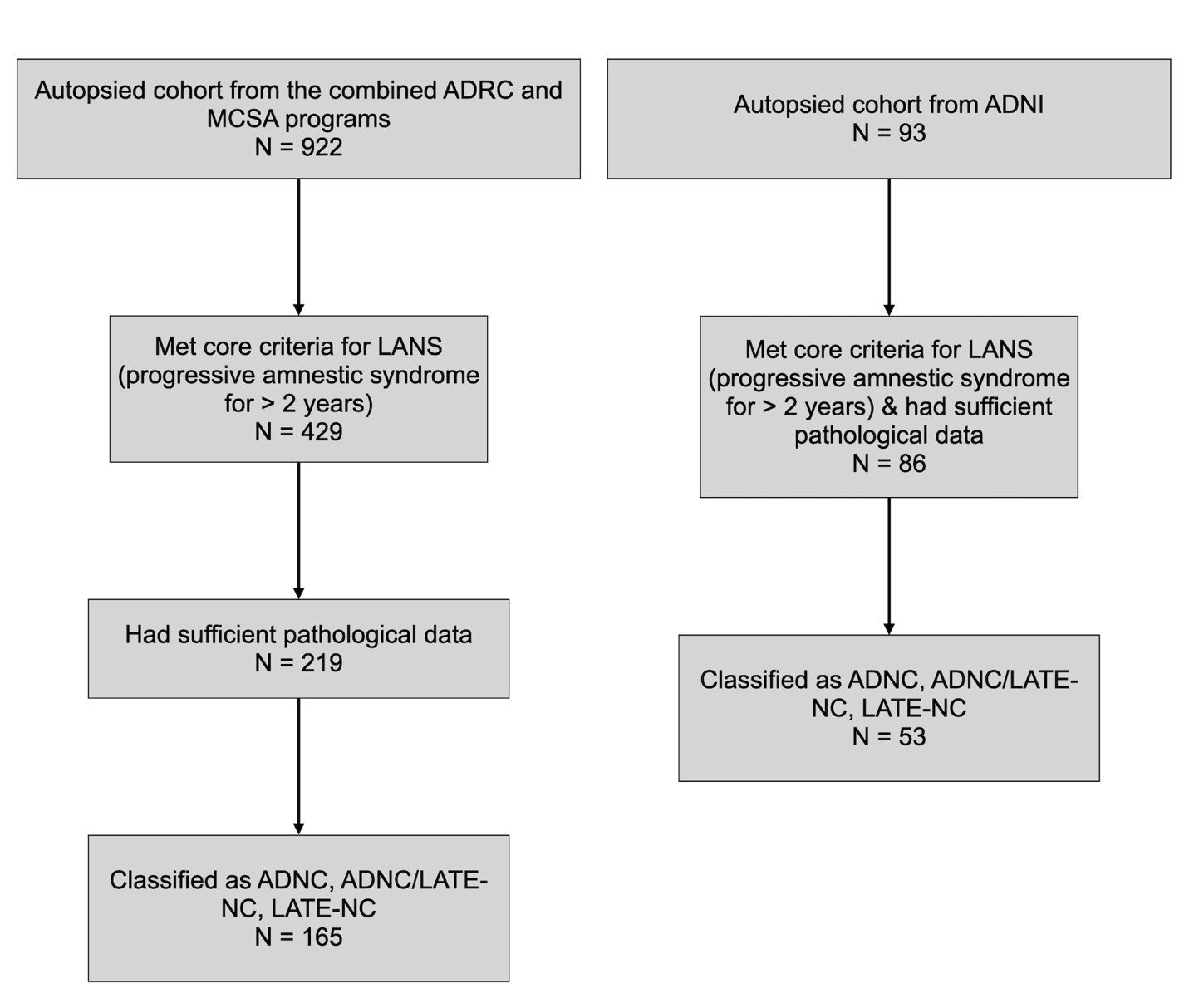

*Supplementary Figure 1. Flow chart of the study.* This Figure displays the selection process of pathological diagnoses cases with a history of progressive and predominant amnestic neurodegenerative syndrome in the Mayo and ADNI cohorts. LANS = Limbic-predominant amnestic neurodegenerative syndrome. Of note, all ADNI who met core criteria for LANS had sufficient pathological data for analysis. ADNC = Alzheimer’s disease neuropathological change; LATE-NC = Limbic-predominant age-associated TDP-43 encephalopathy pathological change.

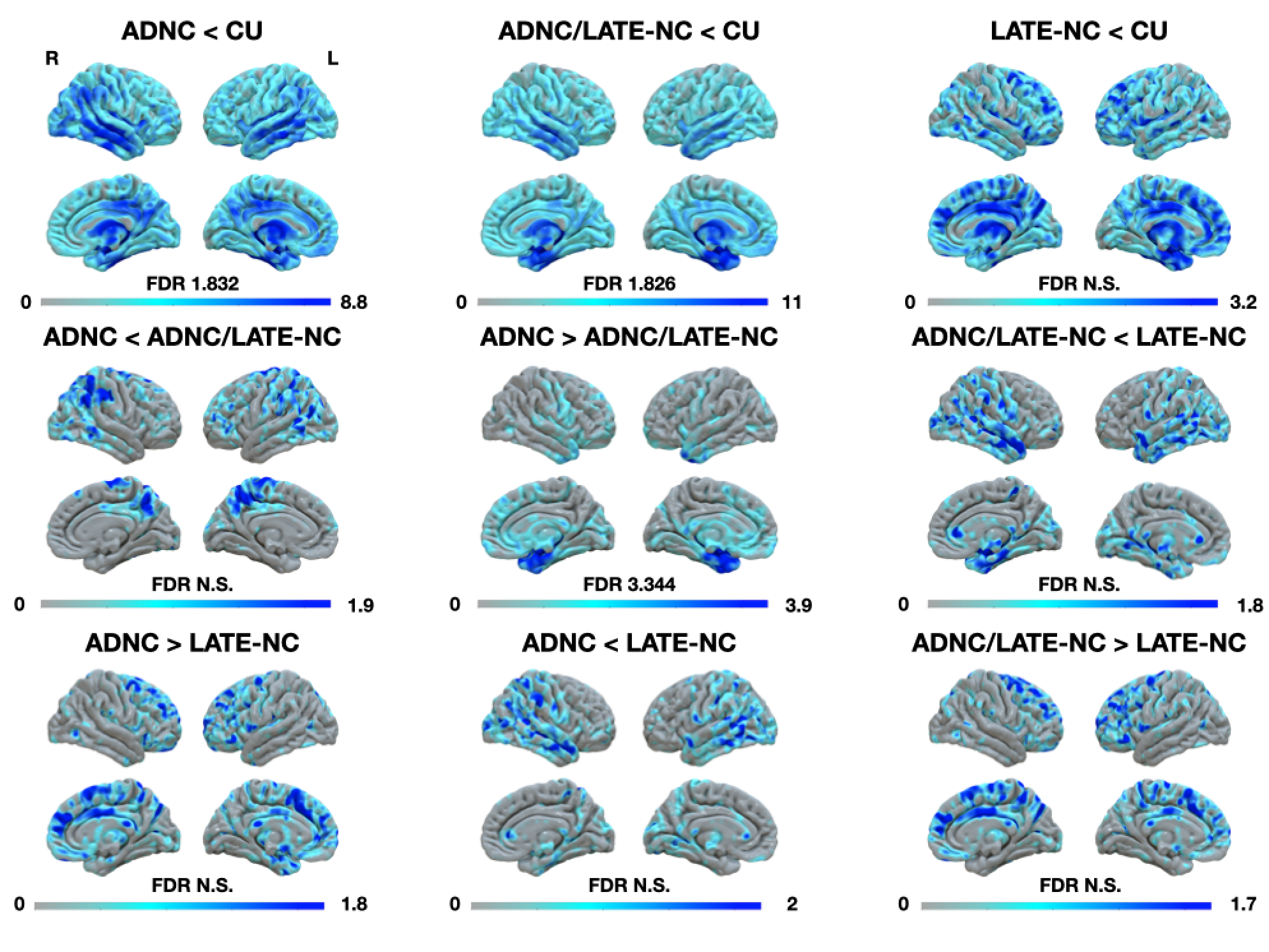

*Supplementary Figure 2. MRI findings between ADNC, ADNC/LATE-NC, LATE-NC, and CU controls.* The “minus than” sign reflects less volume in a given group relative to the other, and vice-versa. ADNC = Alzheimer’s disease neuropathological change; LATE-NC = Limbic-predominant age-associated TDP-43 encephalopathy neuropathological change; CU = Cognitively unimpaired; FDR = False discovery rate; N.S. = Non-significant.

| **Supplementary Table 1 Demographic, clinical, and biomarker data of the Mayo and ADNI cohorts** | | | | | | | | | | | | | | | | | |
| --- | --- | --- | --- | --- | --- | --- | --- | --- | --- | --- | --- | --- | --- | --- | --- | --- | --- |
| Mayo cohort | | | | | | | | | | | | | | | | | |
|  | ADNC | AD-NC/LATE-NC | | LATE-NC | ADNC/LBD | | AGD | AGD/LATE-NC | | FTLD-Tau | FTLD-TDP | | LBD | None | | PART | VaD |
| n | 75 | 81 | | 9 | 30 | | 2 | 2 | | 4 | 4 | | 4 | 1 | | 1 | 5 |
| Age at first visit | 74.2 [63.7, 80.5] | 79 [72.2, 83.9] | | 86.2 [83.6, 93.3] | 71.8 [67.7, 78.4] | | 84.2 [84.1, 84.2] | 86.2 [84.8, 87.6] | | 82.1 [77.6, 86.8] | 78.8 [72.7, 83.9] | | 82.7 [79, 86.1] | 93.9 | | 93.9 | 79.6 [74.5, 81.8] |
| Age at death | 82.4 [71.1, 88.3] | 89.1 [83.9, 92.4] | | 91.6 [88.7, 95.5] | 79.1 [71.5, 90.3] | | 86.4 [86.4, 86.4] | 88.1 [86.7, 89.4] | | 88.6 [85.9, 89.2] | 88.8 [84.8, 92] | | 86.2 [82.8, 90.3] | 95.9 | | 95.4 | 87.2 [83.5, 87.2] |
| Sex (F, M) | 33, 42 | 36, 45 | | 4, 5 | 18, 12 | | 2, 0 | 0, 2 | | 0, 4 | 2, 2 | | 0, 4 | 1, 0 | | 1, 0 | 2, 3 |
| Education | 16 [12,18] | 16 [12, 17] | | 14 [13, 16] | 14 [12, 16.8] | | 12.5 [11.2, 13.8] | 14.5 [13.8, 15.2] | | 12 [12, 13] | 18.5 [17.2, 19.2] | | 12 [11, 13] | 12 | | 12 | 16 [14, 18] |
| CDR-SB | 1.5 [0.5, 3.5] | 1.5 [0.5, 3] | | 0.5 [0.5, 1.25] | 1.75 [1, 4.25] | | 0.5 [0.25, 0.75] | 0.75 [0.63, 0.88] | | 3 [1.38, 5.12] | 1.25 [0.5, 2.38] | | 1 [0.75, 1.5] | 1 | | 0 | 2.5 [0.5, 3] |
| Clinical AD | 27 | 30 | | 1 | 12 | | 0 | 0 | | 2 | 1 | | 0 | 0 | | 0 | 2 |
| Amnestic MCI | 48 | 51 | | 8 | 49 | | 2 | 2 | | 2 | 3 | | 4 | 1 | | 1 | 3 |
| APOE4 (+, -) | 47, 27 | 55, 24 | | 1, 7 | 24, 6 | | 0, 2 | 1, 1 | | 0, 3 | 2, 2 | | 0, 4 | 0, 1 | | 0, 1 | 1, 4 |
| Amyloid-PET centiloid | 110 [87.3, 129] | 110 [76.7, 122] | | 9.75 [7.60, 13.4] | 102 [57.5, 137] | | NA | 62.8* | | 25 [19.9, 30.1] | 131* | | 10.6* | NA | | NA | NA |
| AV1451 SUVr | 1.85 [1.72, 2.28] | 2.14 [1.40, 2.36] | | 1.26* | 1.64 [1.31, 2.03] | | NA | NA | | NA | NA | | NA | NA | | NA | NA |
| ADNI cohort | | | | | | |  |  | |  |  | |  |  | |  |  |
|  | ADNC | | ADNC/LATE-NC | | | LATE | | | ADNC/LBD | | | LBD | | | PART | | |
| n | 26 | | 19 | | | 8 | | | 20 | | | 7 | | | 6 | | |
| Age at first visit | 75.8 [70.3, 81.1] | | 77.7 [72.2, 85.5] | | | 74.9 [72.6, 81.7] | | | 75.9 [72.4, 79.] | | | 81.5 [79.7, 84.1] | | | 79.1 [76, 83.8] | | |
| Age at death | 79 [76.2, 85.8] | | 84 [79, 89] | | | 80.5 [79.8, 87] | | | 81.5 [77.8, 84.5] | | | 88 [84.5, 93] | | | 86 [84.2, 88.5] | | |
| Sex (F, M) | 9, 17 | | 5, 14 | | | 1, 7 | | | 3, 17 | | | 1, 6 | | | 1, 5 | | |
| Education | 16 [16, 18] | | 16 [13.5, 17] | | | 16 [14.5, 18] | | | 16 [16, 19.2] | | | 17 [12, 18.5] | | | 18 [15.8, 19.5] | | |
| CDR-SB | 2 [1, 3.38] | | 3.5 [2, 4.5] | | | 1 [0.875, 1.12] | | | 3 [2.5, 4.5] | | | 2 [1.5, 3] | | | 1.75 [1.12, 3.28] | | |
| Clinical AD | 9 | | 11 | | | 0 | | | 11 | | | 1 | | | 0 | | |
| Amnestic MCI | 17 | | 8 | | | 8 | | | 9 | | | 6 | | | 6 | | |
| APOE4 (+, -) | 20, 6 | | 13, 6 | | | 1, 7 | | | 16, 4 | | | 0, 7 | | | 3, 3 | | |
| Amyloid-PET centiloid | 69.5 [50.2, 110] | | 91 [81, 102] | | | -5 [-11.8, 0] | | | 91.5 [78, 105] | | | -3 [-10, 32] | | | 51 [24.5, 57.5] | | |
| AV1451 SUVr | NA | | 1.52 [1.49, 1.55] | | | 1.08* | | | 1.21* | | | NA | | | 1.19 [1.15, 1.24] | | |
| *One observation only. Values are expressed as counts or median and interquartile ranges. CDR-SB = Clinical Dementia Rating scale – Sum of Boxes; AD = Alzheimer’s disease; ADNC = Alzheimer’s disease neuropathological change; LATE-NC = Limbic-predominant age-related TDP-43 encephalopathy neuropathological change; LBD = Lewy Body disease; PART = Primary age-related tauopathy; MCI = Mild cognitive impairment; PET = Positron emission tomography; SUVr = Standard uptake value ratio. | | | | | | | | | | | | | | | | | |

| **Supplementary Table 2 Vascular burden across pathological diagnoses** | | | | | | | |
| --- | --- | --- | --- | --- | --- | --- | --- |
| Cerebral amyloid angiopathy | | | | | | | |
| Mayo cohort | | | | | | | |
| Diagnosis | None | Mild | | Moderate | Severe | | Unknown |
| ADNC | 7 | 17 | | 37 | 14 | | 1 |
| ADNC/LATE-NC | 4 | 15 | | 40 | 19 | | 6 |
| LATE-NC | 5 | 1 | | 3 | 0 | | 0 |
| ADNC/LBD | 4 | 6 | | 16 | 4 | | 0 |
| AGD | 1 | 0 | | 1 | 0 | | 0 |
| AGD/LATE-NC | 1 | 0 | | 1 | 0 | | 0 |
| FTLD-TDP-43 | 1 | 0 | | 3 | 0 | | 0 |
| FTLD-Tau | 1 | 0 | | 3 | 0 | | 0 |
| LBD | 1 | 1 | | 2 | 0 | | 0 |
| None | 0 | 1 | | 0 | 0 | | 0 |
| PART | 1 | 0 | | 0 | 0 | | 0 |
| VaD | 2 | 2 | | 1 | 0 | | 0 |
| ADNI cohort | | | | | | | |
| ADNC | 0 | 12 | | 7 | 7 | | 0 |
| ADNC/LATE-NC | 2 | 10 | | 4 | 3 | | 0 |
| LATE-NC | 1 | 5 | | 1 | 1 | | 0 |
| ADNC/LBD | 1 | 7 | | 6 | 6 | | 0 |
| LBD | 4 | 1 | | 2 | 0 | | 0 |
| PART | 3 | 3 | | 0 | 0 | | 0 |
| Infarcts including lacunes | | | | | | | |
| Mayo cohort | | | | | | | |
| Diagnosis | Present | | Absent | | | Unknown | |
| ADNC | 9 | | 20 | | | 47 | |
| ADNC/LATE-NC | 9 | | 34 | | | 38 | |
| LATE-NC | 5 | | 2 | | | 2 | |
| ADNC/LBD | 2 | | 17 | | | 11 | |
| AGD | 1 | | 1 | | | 0 | |
| AGD/LATE-NC | 0 | | 1 | | | 1 | |
| FTLD-TDP-43 | 1 | | 3 | | | 0 | |
| FTLD-Tau | 0 | | 3 | | | 1 | |
| LBD | 1 | | 2 | | | 1 | |
| None | 0 | | 0 | | | 1 | |
| PART | 1 | | 0 | | | 0 | |
| VaD | 3 | | 2 | | | 0 | |
| ADNI cohort | | | | | | | |
| ADNC | 2 | | 24 | | | 0 | |
| ADNC/LATE-NC | 2 | | 17 | | | 0 | |
| LATE-NC | 0 | | 8 | | | 0 | |
| ADNC/LBD | 2 | | 17 | | | 0 | |
| LBD | 7 | | 0 | | | 0 | |
| PART | 1 | | 5 | | | 0 | |
| Microbleeds | | | | | | | |
| Mayo cohort | | | | | | | |
| Diagnosis | Present | | Absent | | | Unknown | |
| ADNC | 8 | | 19 | | | 49 | |
| ADNC/LATE-NC | 6 | | 34 | | | 41 | |
| LATE-NC | 1 | | 1 | | | 7 | |
| ADNC/LBD | 1 | | 18 | | | 11 | |
| AGD | 2 | | 0 | | | 0 | |
| AGD/LATE-NC | 0 | | 1 | | | 1 | |
| FTLD-TDP-43 | 0 | | 3 | | | 1 | |
| FTLD-Tau | 0 | | 3 | | | 1 | |
| LBD | 1 | | 2 | | | 1 | |
| None | 0 | | 0 | | | 1 | |
| PART | 0 | | 0 | | | 1 | |
| VaD | 1 | | 3 | | | 1 | |
| ADNI cohort | | | | | | | |
| ADNC | 0 | | 26 | | | 0 | |
| ADNC/LATE-NC | 0 | | 19 | | | 0 | |
| LATE-NC | 0 | | 8 | | | 0 | |
| ADNC/LBD | 0 | | 20 | | | 0 | |
| LBD | 0 | | 7 | | | 0 | |
| PART | 0 | | 6 | | | 0 | |
| Hemorrhages | | | | | | | |
| Mayo cohort | | | | | | | |
| Diagnosis | Present | | Absent | | | Unknown | |
| ADNC | 4 | | 21 | | | 51 | |
| ADNC/LATE-NC | 5 | | 36 | | | 40 | |
| LATE-NC | 0 | | 1 | | | 8 | |
| ADNC/LBD | 1 | | 17 | | | 12 | |
| AGD | 0 | | 2 | | | 0 | |
| AGD/LATE-NC | 0 | | 1 | | | 1 | |
| FTLD-TDP-43 | 0 | | 3 | | | 1 | |
| FTLD-Tau | 0 | | 3 | | | 1 | |
| LBD | 0 | | 3 | | | 1 | |
| None | 0 | | 0 | | | 1 | |
| PART | 0 | | 0 | | | 1 | |
| VaD | 0 | | 4 | | | 1 | |
| ADNI cohort | | | | | | | |
| ADNC | 2 | | 24 | | | 0 | |
| ADNC/LATE-NC | 1 | | 18 | | | 0 | |
| LATE-NC | 0 | | 8 | | | 0 | |
| ADNC/LBD | 1 | | 18 | | | 0 | |
| LBD | 0 | | 7 | | | 0 | |
| PART | 0 | | 6 | | | 0 | |
| Arteriolosclerosis | | | | | | | |
| Mayo cohort | | | | | | | |
| Diagnosis | None | Mild | | Moderate | Severe | | Unknown |
| ADNC | 6 | 12 | | 28 | 27 | | 3 |
| ADNC/LATE-NC | 2 | 5 | | 40 | 28 | | 6 |
| LATE-NC | 0 | 3 | | 4 | 2 | | 0 |
| ADNC/LBD | 4 | 2 | | 12 | 10 | | 2 |
| AGD | 1 | 0 | | 0 | 1 | | 0 |
| AGD/LATE-NC | 1 | 1 | | 0 | 0 | | 0 |
| FTLD-TDP-43 | 1 | 0 | | 2 | 1 | | 0 |
| FTLD-Tau | 0 | 0 | | 3 | 0 | | 1 |
| LBD | 0 | 0 | | 0 | 3 | | 1 |
| None | 0 | 1 | | 0 | 0 | | 0 |
| PART | 0 | 0 | | 1 | 0 | | 0 |
| VaD | 0 | 1 | | 1 | 3 | | 0 |
| ADNI cohort | | | | | | | |
| ADNC | 1 | 16 | | 7 | 2 | | 0 |
| ADNC/LATE-NC | 1 | 13 | | 2 | 3 | | 0 |
| LATE-NC | 0 | 5 | | 2 | 1 | | 0 |
| ADNC/LBD | 1 | 10 | | 8 | 1 | | 0 |
| LBD | 0 | 5 | | 5 | 0 | | 0 |
| PART | 0 | 4 | | 4 | 0 | | 0 |
| ADNC = Alzheimer’s disease neuropathological change; LATE-NC = Limbic-predominant age-associated TDP-43 encephalopathy neuropathological change; LBD = Lewy Body disease; AGD = Argyrophilic grain disease; FTLD = Fronto-temporal lobar degeneration; PART = Primary age-related tauopathy; VaD = Vascular disease. | | | | | | | |

| **Supplementary Table 3 LANS features and likelihoods across pathological diagnoses for the Mayo and ADNI cohorts** | | | | | | | | | | | | | | |
| --- | --- | --- | --- | --- | --- | --- | --- | --- | --- | --- | --- | --- | --- | --- |
| Mayo cohort | | | | | | | | | | | | | | |
| Standard and advanced LANS features (meets feature/total) | | | | | | | | | | | | | | |
| Diagnosis | Age ≥75 | Mild syndrome | | | Hippocampal atrophy | | Limbic hypometabolism | | | Absence of neocortical hypometabolism | | | Low neocortical tau likelihood | |
| ADNC | 36/75 | 58/75 | | | 25/63 | | 9/53 | | | 5/53 | | | 7/60 | |
| ADNC/LATE-NC | 53/81 | 66/81 | | | 38/54 | | 15/33 | | | 7/33 | | | 1/49 | |
| LATE-NC | 8/9 | 8/9 | | | 3/4 | | 3/4 | | | 1/4 | | | 5/7 | |
| ADNC/LBD | 11/30 | 22/30 | | | 7/19 | | 0/11 | | | 1/11 | | | 2/19 | |
| LBD | 4/4 | 4/4 | | | 0/1 | | 0/2 | | | 0/2 | | | 2/2 | |
| AGD | 2/2 | 2/2 | | | NA | | NA | | | NA | | | 0/2 | |
| AGD/LATE-NC | 2/2 | 2/2 | | | 0/1 | | 1/1 | | | 1/1 | | | 0/1 | |
| FTLD-TDP-43 | 2/4 | 4/4 | | | 3/4 | | 1/1 | | | 0/1 | | | 0/2 | |
| FTLD-Tau | 4/4 | 2/2 | | | 0/2 | | 1/2 | | | 1/2 | | | 1/3 | |
| PART | 1/1 | 1/1 | | | NA | | NA | | | NA | | | 1/1 | |
| Vascular | 3/5 | 5/5 | | | 0/2 | | NA | | | NA | | | NA | |
| None | 1/1 | 1/1 | | | NA | | NA | | | NA | | | NA | |
| LANS likelihoods (only including patients with all features available) | | | | | | | | | | | | | | |
| Diagnosis | Low | | | Moderate | | | | | High | | | Highest | | |
| ADNC | 31 | | | 17 | | | | | 1 | | | 0 | | |
| ADNC/LATE-NC | 12 | | | 8 | | | | | 13 | | | 0 | | |
| LATE-NC | 0 | | | 0 | | | | | 2 | | | 2 | | |
| ADNC/LBD | 9 | | | 2 | | | | | 0 | | | 0 | | |
| LBD | 0 | | | 1 | | | | | 0 | | | 0 | | |
| AGD/LATE-NC | 0 | | | 0 | | | | | 1 | | | 0 | | |
| FTLD-TDP-43 | 0 | | | 1 | | | | | 0 | | | 0 | | |
| FTLD-Tau | 1 | | | 0 | | | | | 1 | | | 0 | | |
| ADNI cohort | | | | | | | | | | | | | | |
| Standard and advanced LANS features | | | | | | | | | | | | | | |
| Diagnosis | Age ≥75 | | Mild syndrome | | | Hippocampal atrophy | | Limbic hypometabolism | | | Absence of neocortical hypometabolism | | | Low neocortical tau likelihood |
| ADNC | 15/26 | | 20/26 | | | 11/26 | | 1/18 | | | 2/18 | | | 1/22 |
| ADNC/LATE-NC | 12/19 | | 12/19 | | | 15/19 | | 4/13 | | | 1/13 | | | 0/17 |
| LATE-NC | 4/8 | | 8/8 | | | 5/8 | | 5/6 | | | 1/6 | | | 8/8 |
| ADNC/LBD | 11/20 | | 14/20 | | | 12/20 | | 5/16 | | | 0/16 | | | 0/17 |
| LBD | 7/7 | | 6/7 | | | 3/6 | | 0/4 | | | 0/4 | | | 2/5 |
| PART | 5/6 | | 6/6 | | | 1/6 | | 1/5 | | | 2/5 | | | 4/6 |
| LANS likelihoods (only including patients with all features available) | | | | | | | | | | | | | | |
| Diagnosis | Low | | | Moderate | | | | | High | | | Highest | | |
| ADNC | 11 | | | 7 | | | | | 0 | | | 0 | | |
| ADNC/LATE-NC | 8 | | | 3 | | | | | 1 | | | 0 | | |
| LATE-NC | 2 | | | 0 | | | | | 1 | | | 3 | | |
| ADNC-LBD | 8 | | | 4 | | | | | 3 | | | 0 | | |
| LBD | 2 | | | 0 | | | | | 2 | | | 0 | | |
| PART | 2 | | | 1 | | | | | 1 | | | 1 | | |
| LANS = Limbic-predominant amnestic neurodegenerative syndrome; ADNC = Alzheimer’s disease neuropathological changes; LATE-NC = Limbic-predominant age-related TDP-43 encephalopathy neuropathological changes; LBD = Lewy Body disease; FTLD = Frontotemporal lobar degeneration; AGD = Argyrophilic grain disease; PART = Primary age-related tauopathy. | | | | | | | | | | | | | | |

| **Supplementary Table4 Mixed linear modelling results between likelihood categories and longitudinal CDR-SB scores since baseline** | | | | | | |
| --- | --- | --- | --- | --- | --- | --- |
| Baseline comparisons | | | | | | |
| Within-groups divergences from intercept (i.e., CDR-SB of 0) | | | | | | |
|  | Highest | High | | Moderate | | Low |
| Estimated marginal mean | 0.79 | 2.25 | | 1.25 | | 2.95 |
| Standard error | 2.36 | 0.84 | | 0.68 | | 0.51 |
| DF | 104 | 105 | | 108 | | 106 |
| Confidence intervals | -3.88; 5.46 | 0.59; 3.91 | | -0.09; 2.59 | | 1.94; 3.97 |
| Between-groups comparisons at baseline | | | | | | |
| Contrast | Estimate | Standard error | DF | | *T* ratio | *P* value |
| Highest – High | -1.46 | 2.5 | 104 | | -0.58 | 0.94 |
| Highest – Moderate | -0.46 | 2.45 | 104 | | -0.19 | 0.99 |
| Highest – Low | -2.16 | 2.41 | 104 | | -0.90 | 0.81 |
| High – Moderate | 0.99 | 1.08 | 106 | | 0.92 | 0.79 |
| High – Low | -0.70 | 0.98 | 105 | | -0.72 | 0.89 |
| Moderate – Low | -1.70 | 0.85 | 107 | | -2.00 | 0.19 |
| Slopes comparisons | | | | | | |
| Within-groups assessment of slope change | | | | | | |
|  | Highest | High | | Moderate | | Low |
| Estimated marginal trend | -0.12 | 0.40 | | 0.76 | | 1.02 |
| Standard error | 0.48 | 0.11 | | 0.09 | | 0.06 |
| DF | 252 | 287 | | 298 | | 285 |
| Confidence intervals | -1.07; 0.83 | 0.18; 0.62 | | 0.57; 0.94 | | 0.87; 1.17 |
| Between-groups comparisons of slope change | | | | | | |
| Contrast | Estimate | Standard error | DF | | *T* ratio | *P* value |
| Highest – High | -0.52 | 0.497 | 254 | | -1.040 | 0.72 |
| Highest – Moderate | -0.88 | 0.49 | 254 | | -1.78 | 0.29 |
| Highest – Low | -1.14 | 0.49 | 253 | | -2.33 | 0.09 |
| High – Moderate | -0.36 | 0.15 | 292 | | -2.44 | 0.07 |
| High – Low | -0.62 | 0.14 | 287 | | -4.56 | <0.001 |
| Moderate – Low | -0.26 | 0.12 | 293 | | -2.19 | 0.13 |
| CDR-SB = Clinical dementia rating scale – Sum of Boxes; DF = Degrees of freedom. | | | | | | |

| **Supplementary Table 5 Mixed linear modelling results between pathological diagnoses and longitudinal CDR-SB scores since baseline** | | | | | |
| --- | --- | --- | --- | --- | --- |
| Baseline comparisons | | | | | |
| Within-groups divergences from intercept (i.e., CDR-SB of 0) | | | | | |
|  | ADNC | ADNC/LATE-NC (H) | ADNC/LATE-NC (M) | ADNC/LATE-NC (L) | LATE-NC |
| Estimated marginal mean | 2.85 | 2.38 | 1.07 | 2.57 | 1.27 |
| Standard error | 0.40 | 0.94 | 1.21 | 0.97 | 1.21 |
| DF | 135 | 136 | 139 | 130 | 139 |
| Confidence intervals | 2.06; 3.64 | 0.52; 4.25 | -1.32; 3.47 | 0.65; 4.49 | -1.12; 3.66 |
| Between-groups comparisons at baseline | | | | | |
| Contrast | Estimate | Standard error | DF | *T* ratio | *P* value |
| ADNC – ADNC/LATE-HC (H) | 0.46 | 1.02 | 136 | 0.45 | 0.99 |
| ADNC – ADNC/LATE-HC (M) | 1.78 | 1.27 | 138 | 1.39 | 0.63 |
| ADNC – ADNC/LATE-HC (L) | 0.28 | 1.05 | 130 | 0.27 | 0.99 |
| ADNC – LATE-NC | 1.58 | 1.27 | 138 | 1.24 | 0.73 |
| ADNC/LATE-NC (H) – ADNC/LATE-NC (M) | 1.31 | 1.53 | 138 | 0.85 | 0.91 |
| ADNC/LATE-NC (H) – ADNC/LATE-NC (L) | -0.19 | 1.35 | 133 | -0.14 | 1.00 |
| ADNC/LATE-NC (H) – LATE-NC | 1.12 | 1.53 | 138 | 0.73 | 0.95 |
| ADNC/LATE-NC (M) – ADNC/LATE-NC (L) | -1.45 | 1.55 | 135 | -0.97 | 0.87 |
| ADNC/LATE-NC (M) – LATE-NC | -0.20 | 1.71 | 139 | -0.11 | 1.00 |
| ADNC/LATE-NC (L) – LATE-NC | 1.30 | 1.55 | 135 | 0.84 | 0.92 |
| Slopes comparisons | | | | | |
| Within-groups assessment of slope change | | | | | |
|  | ADNC | ADNC/LATE-NC (H) | ADNC/LATE-NC (M) | ADNC/LATE-NC (L) | LATE-NC |
| Estimated marginal trend | 2.85 | 2.38 | 1.07 | 2.57 | 1.27 |
| Standard error | 0.40 | 0.94 | 1.21 | 0.97 | 1.21 |
| DF | 135 | 136 | 139 | 130 | 139 |
| Confidence intervals | 2.06; 3.64 | 0.52; 4.25 | -1.32; 3.47 | 0.65; 4.49 | -1.12; 3.66 |
| Between-groups comparisons of slope change | | | | | |
| Contrast | Estimate | Standard error | DF | *T* ratio | *P* value |
| ADNC – ADNC/LATE-HC (H) | 0.30 | 0.13 | 352 | 2.32 | 0.14 |
| ADNC – ADNC/LATE-HC (M) | 0.03 | 0.17 | 358 | 0.17 | 1.00 |
| ADNC – ADNC/LATE-HC (L) | -0.57 | 0.13 | 341 | -4.28 | <0.001 |
| ADNC – LATE-NC | 0.44 | 0.29 | 353 | 1.54 | 0.54 |
| ADNC/LATE-NC (H) – ADNC/LATE-NC (M) | -0.27 | 0.19 | 355 | -1.48 | 0.58 |
| ADNC/LATE-NC (H) – ADNC/LATE-NC (L) | -0.87 | 0.17 | 343 | -5.21 | <0.001 |
| ADNC/LATE-NC (H) – LATE-NC | 0.14 | 0.31 | 352 | 0.47 | 0.99 |
| ADNC/LATE-NC (M) – ADNC/LATE-NC (L) | -0.60 | 0.19 | 349 | -3.17 | 0.01 |
| ADNC/LATE-NC (M) – LATE-NC | 0.42 | 0.32 | 354 | 1.31 | 0.68 |
| ADNC/LATE-NC (L) – LATE-NC | 1.02 | 0.31 | 350 | 3.30 | 0.01 |
| CDR-SB = Clinical dementia rating scale – Sum of Boxes; DF = Degrees of freedom; ADNC = Alzheimer’s disease neuropathological changes; LATE-NC = Limbic-predominant age-related TDP-43 encephalopathy neuropathological changes; (H) = Highest/high likelihoods; (M) = Moderate likelihood; (L) = Low likelihood. | | | | | |

**References**

1.	Jack CR, Wiste HJ, Weigand SD, Therneau TM, Lowe VJ, Knopman DS, et al. Defining imaging biomarker cut points for brain aging and Alzheimer’s disease. Alzheimer’s Dement. 2017;13(3):205–16.

2.	Jack CR, Knopman DS, Weigand SD, Wiste HJ, Vemuri P, Lowe V, et al. An operational approach to National Institute on Aging-Alzheimer’s Association criteria for preclinical Alzheimer disease. Ann Neurol. 2012;71(6):765–75.

3.	Cairns NJ, Taylor-Reinwald L, Morris JC, Initiative ADN. Autopsy consent, brain collection, and standardized neuropathologic assessment of ADNI participants: the essential role of the neuropathology core. Alzheimer’s Dement. 2010;6(3):274–9.

4.	Toledo JB, Cairns NJ, Da X, Chen K, Carter D, Fleisher A, et al. Clinical and multimodal biomarker correlates of ADNI neuropathological findings. Acta Neuropathol Commun. 2013;1(1):1–13.

5.	Franklin EE, Perrin RJ, Vincent B, Baxter M, Morris JC, Cairns NJ, et al. Brain collection, standardized neuropathologic assessment, and comorbidity in Alzheimer’s Disease Neuroimaging Initiative 2 participants. Alzheimer’s Dement. 2015;11(7):815–22.
